# A frequent ancestral NFKB1 variant predicts risk of infection or allergy

**DOI:** 10.1101/2022.11.24.22282707

**Authors:** A. Y. Chong, N. Brenner, A. Jimenez-Kaufmann, A. Cortes, M. Hill, T. J. Littlejohns, J. J. Gilchrist, B. P. Fairfax, J. C. Knight, F. Hodel, J. Fellay, G. McVean, A. Moreno-Estrada, T. Waterboer, A. V. S. Hill, A. J. Mentzer

## Abstract

Infectious agents contribute significantly to the global burden of diseases, through both acute infection and their chronic sequelae. We leveraged the UK Biobank to identify genetic loci that influence humoral immune response to multiple infections. From 45 genome-wide association studies in 9,611 participants from UK Biobank, we identified *NFKB1* as a locus associated with quantitative antibody responses to multiple pathogens including those from the herpes, retro- and polyoma-virus families. An insertion-deletion variant thought to affect *NFKB1* expression (rs28362491), was mapped as the likely causal variant. This variant has persisted throughout hominid evolution and could play a key role in regulation of the immune response. Using 121 infection and inflammation related traits in 487,297 UK Biobank participants, we show that the deletion allele was associated with an increased risk of infection from diverse pathogens but had a protective effect against allergic disease. We propose that altered expression of *NFKB1*, as a result of the deletion, modulates haematopoietic pathways, and likely impacts cell survival, antibody production, and inflammation. Taken together, we show that disruptions to the tightly regulated immune processes may tip the balance between exacerbated immune responses and allergy, or increased risk of infection and impaired resolution of inflammation.

## Main text

There is significant evidence that human genetic variation influences susceptibility to acute disease caused by infectious pathogens. Case-control genome-wide association studies (GWAS) of a variety of infections such as human immunodeficiency virus, hepatitis B and C viruses, leprosy, typhoidal and non-typhoidal *Salmonella*, meningococcal disease, and more recently SARS-CoV-2 have identified a range of variants spanning pathogen recognition, immune activation and immunologic memory robustly associated with differential susceptibility ^1–10^. Furthermore, using large biobanks of human genetic data linked with participant self-report, studies such as 23andMe, are replicating and building on these findings from case-control analyses of infection susceptibility ^11^. Together, these studies are helping improve our understanding of how these altered responses to infection may influence our susceptibility to future sequelae of infection such as meningitis, mononucleosis, plantar warts and rheumatic heart disease. Despite these advances, there remain challenges to using both study approaches. Firstly, achieving sufficient power to identify novel genetic signals in case-control studies is limited by challenges in recruiting large numbers of cases, and both study approaches may be affected by pathogen heterogeneity that may impact on signatures of genetic association, whilst the self-report approach will be influenced by limitations in accuracy and recall of self-reporting.

Antibodies are a critical component of response to infection and represent a marker of prior exposure or chronic carriage with agents capable of latent or chronic infection states. Now that methods are available to accurately measure multiple antibodies simultaneously in large numbers of individuals, these represent stable biological markers that can be used to understand disease risk. Furthermore, there is evidence from vaccine studies that the magnitude of antibody response and levels of circulating antibody may reflect the likelihood of protection against either primary or subsequent disease, although this may depend on the antigen to which the antibody is directed, and the functional impact of antibody binding and activity ^12^.

Human genetic variation is recognised to significantly influence total antibody levels ^13^. Genetic analyses of antibody response levels against vaccine preventable infections, such as hepatitis B virus, have demonstrated a strong influence of genetic loci, including the major histocompatibility locus (MHC) in predicting antibody magnitude overlapping with GWAS of disease susceptibility ^14^. The MHC has also been implicated in varied antibody responses to infections including JC virus, and influenza ^15–17^. Furthermore, primary immunodeficiencies such as common variable immunodeficiency (CVID) and X-linked agammaglobulinaemia are well recognised disorders that increase the risk of multiple infections owing to impaired humoral function and antibody deficiencies ^18–20^.

The availability of antibody data against 45 antigens representing 20 infectious agents in combination with genome-wide genotyping data in a subset of UK Biobank participants now offers the opportunity to investigate whether there are common variants that influence both response to antibodies and subsequent susceptibility to both acute infectious disease and their chronic sequelae. While previous studies have identified genetic associations with single antigens or pathogens using this UK Biobank dataset ^17,21^, we hypothesise that there are genetic variants and loci that have a common effect on antibody responses to multiple infections through effects on central immunological pathways. To identify genetic loci that influence antibody titres against multiple infections, we performed GWAS for magnitude of antibody responses to each antigen and identified *NFKB1* as a novel and distinct locus of interest, on contrast to the other, more familiar, loci that were identified including immunoglobulin genes, the major histocompatibility complex and *FUT2*. Given the role of *NFKB1* in inflammation and the induction and regulation of immune responses to infection, we further investigated the association between *NFKB1* variation and infectious and inflammatory diseases using data from 487,297 individuals. Finally, to better understand the impact of variation in this locus on underlying immune mechanisms, we explored the effect of this variant on blood cell traits and cellular expression data both within UK Biobank and in dedicated expression quantitative trait datasets.

### *NFKB1* is associated with antibody responses against multiple pathogens

We have previously described the use of a Multiplex Serology panel to measure immunoglobulin G (IgG) antibody responses against 45 antigens from 20 infectious agents implicated in the pathogenesis of chronic disease, applied to 9,695 individuals in UK Biobank (^22^; Supplementary Tables 1 and 2 of this manuscript). Although we observed that antibody responses to antigens from the same pathogen were more strongly correlated with each other than with those of other pathogens (Supplementary Fig. 1), we did observe all antibody responses to be weakly correlated with each other (Spearman‘s rho -0.05-0.86). It is likely that shared genetic variants and environmental factors influence these correlated responses. Using the 9,611 individuals with both serology data and imputed genetic data (Supplementary Tables 1 and 2), we performed genome-wide association studies of quantitative antibody responses to 45 antigens to identify these common genetic loci that influence response to multiple infections.

We identified 27 genome-wide significant (*P* < 5×10^−8^) loci associated with magnitude of antibody responses to any antigen (Fig. 1; Extended data Table 1). We observed four genomic regions, localising to *NFKB1* (most significant association with HHV-6 (IE1A); beta = -0.10; *P* = 1.30×10^−10^), the extended MHC region (EBV (EBNA1); beta = -0.32; *P* = 6.90×10^−97^) the IGH locus (HPV16 (E6); beta = 0.19; *P* = 1.60×10^−10^), and *FUT2* (JCV (VP1); beta = -0.13; *P* = 1.70×10^−21^), which were associated with antibody response to multiple antigens, and where at least one association signal passed Bonferroni correction (*P* < 1.1×10^−9^). While all four genetic regions are well recognised to drive differences in infection susceptibility, this is the first GWAS to identify *NFKB1* in relation to antibody responses against infectious disease ^6,11,14–17,21,23^.

**Fig. 1:**
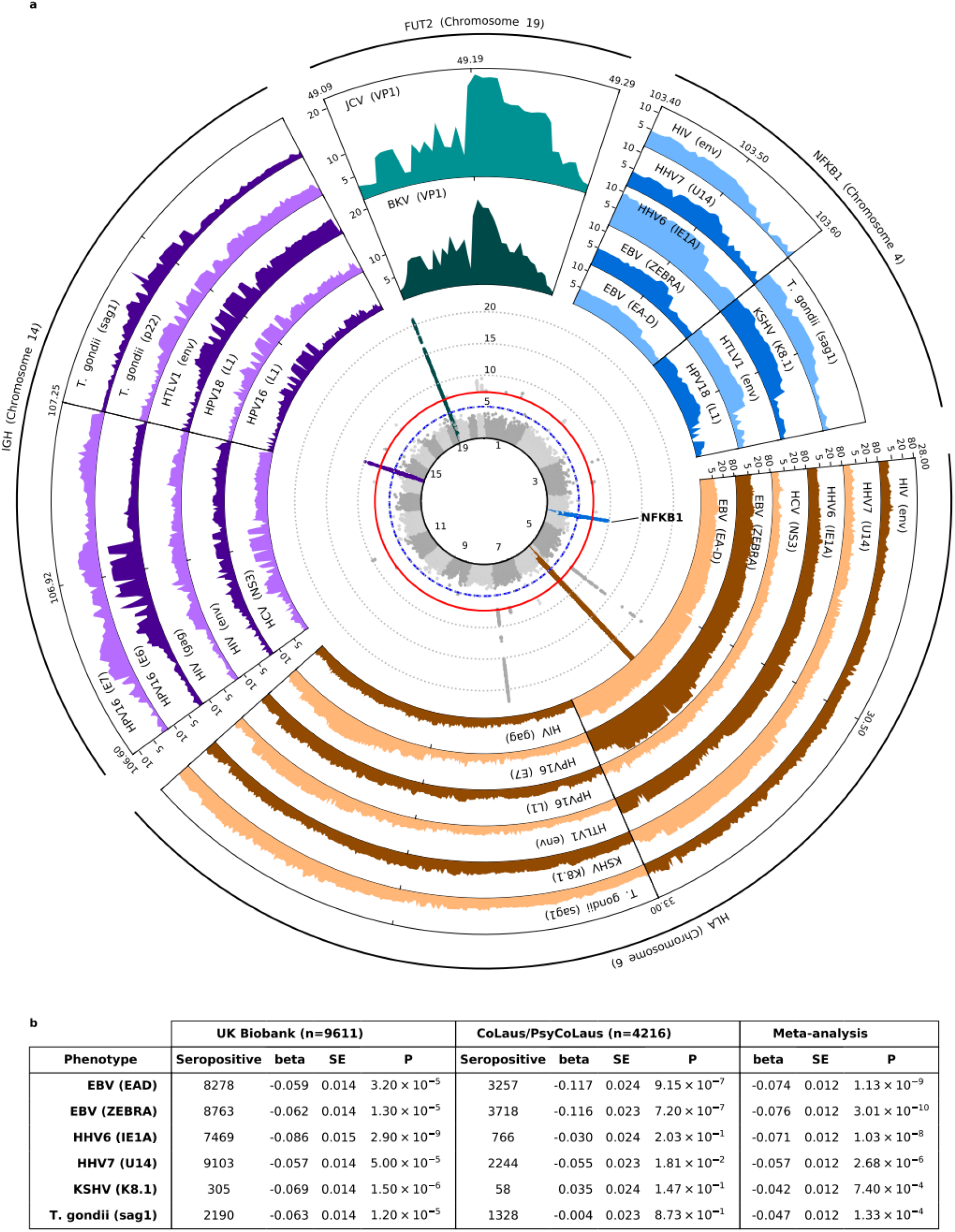
*NFKB1* is a key locus affecting antibody response, and risk of infection and inflammatory disease. (a) Central inset: stacked Manhattan plot of association statistics for antibody response to 45 pathogen-derived antigens. Signals for the four loci with multiple associations are coloured: *NFKB1* (blue), the MHC locus (brown), the IGH locus (purple), and *FUT2* (teal). Surrounding insets: Regional association plots for specific antigens demonstrating signals at each of the four highlighted loci. Y-axis (all plots) is –log_10_(P) (b) Meta-analysis of the NFKB1 locus for association with six antigens in common between the UK Biobank and CoLaus/PsyCoLaus cohorts. Results for rs28362491 are presented.

Associations were observed within the extended MHC region for at least one antigen from each of the 20 pathogens, with associations for human cytomegalovirus (CMV), *Chlamydia trachomatis*, Epstein-Barr virus (EBV), human herpesvirus 6 and 7 (HHV-6 and 7), human papillomavirus type 16 (HPV16), herpes simplex virus types 1 and 2 (HSV-1 and -2), human T-lymphotropic virus 1 (HTLV-1), JC polyomavirus (JCV), Merkel cell polyomavirus (MCV), *Toxoplasma gondii* (*T. gondii*), and varicella-zoster virus (VZV) reaching genome-wide significance (Extended Data Table 2). This suggests that, with adequate power, we are now able to resolve HLA associations with responses to antigens considered most clinically relevant (such as EBNA-1 and EA for EBV; Fig. 1a inset), such as those described from the same UK Biobank cohort ^16,17,21^.

Of particular interest is the identification of multiple variants in *NFKB1* as a common locus associated with antibody responses across multiple pathogens and pathogen types, including EBV, human papillomavirus type 18 (HPV18), HIV, HTLV-1, Kaposi’s sarcoma-associated herpesvirus (KSHV), and *T. gondii*. To confirm the likely contribution of this locus to multiple infectious antigen responses we replicated this association demonstrating a consistent direction of effect for three antigens (EBV EAD, EBV ZEBRA and HHV-7 U14) using an independently recruited cohort (CoLaus/PsyCoLaus; see Supplementary Table 2, Extended Data Table 4, ^16^) where data using the same Multiplex Serology technology was available but with a different set of antigens (Fig. 1b). *NFKB1* is one of five genes that encode the NF-κB family of transcription factors. The NF-κB family plays a critical role in induction and mediation of pro-inflammatory response to infection, and for development and maintenance of blood cells, and immune tissues through modulation of apoptosis. Transcriptomic profiling of immune response to pathogens suggests that *NFKB1* forms part of a larger set of genes involved in regulation of immune response to a wide range of pathogen infections ^24^. Furthermore, mutations in *NFKB1* have been identified as one of the most common causes of monogenic CVID in European populations ^18,20^.

### Ancestral variation in *NFKB1* is under balancing selection in humans and may have conferred evolutionary benefits in early hominids

Using a combination of fine mapping and examination of possible variant effects, we identified the most likely causative variant to be rs28362491, a 4bp insertion-deletion (indel) variant within a promoter region upstream of *NFKB1* which has been associated with multiple disease phenotypes in a range of previously published genetic association studies. Examples of such traits with significant evidence of effect include coronary artery disease with consistent reports of increased odds of disease with presence of the deletion demonstrated in European (Odds ratio (OR) = 2.88 (95% CI 1.21-6.84), Extended Data Table 3), Indian (OR = 1.26 (1.03-1.55)) and Uyghur (OR = 1.58 (1.22-2.05)) populations; risk of lung injury in acute respiratory distress syndrome in Europeans (OR = 3.7 (1.8-7.9)); and ulcerative colitis in Europeans (OR = 1.57 (1.14-2.16)). Furthermore, the same region has been found to be associated in the same direction for other traits where although the rs28362491 variant was not reported, it is likely that the effect may co-localise. Traits of interest include tonsillectomy (OR = 1.08 (1.06-1.08)), ankylosing spondylitis (OR = 1.12 (1.08-1.16)), primary biliary cirrhosis (OR = 1.34 (1.11-1.23)) and mouth ulcers (OR = 1.03 (1.02-1.04)) amongst others (Extended Data Table 3). Intriguingly the same locus and/or variant was associated with other traits in the opposite direction including allergic rhinitis (OR = 0.96 (0.95-0.97)), hayfever and eczema (OR = 0.96 (0.95-0.97)) and cancer risk (OR = 0.89 (0.83-0.96)). The rs28362491 deletion is proposed to disrupt a promoter binding site, lowering *NFKB1* expression ^25^. Given the broad range of effects of this allele and locus on diverse traits in worldwide populations we explored global population frequencies of the deletion allele (rs28362491:delATTG) using data from the 1000 Genomes Project and found evidence using Tajima’s D test for non-neutrality ^26,27^ that variation within this locus is maintained through balancing selection, (Fig. 2b,c). Notably, Tajima’s D was negative for West Africans (YRI, D = -1.34 --0.81), where allele frequency of the deletion allele was higher than global average, than for other populations (CEU: D = 0.37-1.58, CHB: D = 0.67 - 2.05) suggesting that this locus is still undergoing purifying selection in some population groups.

**Fig. 2:**
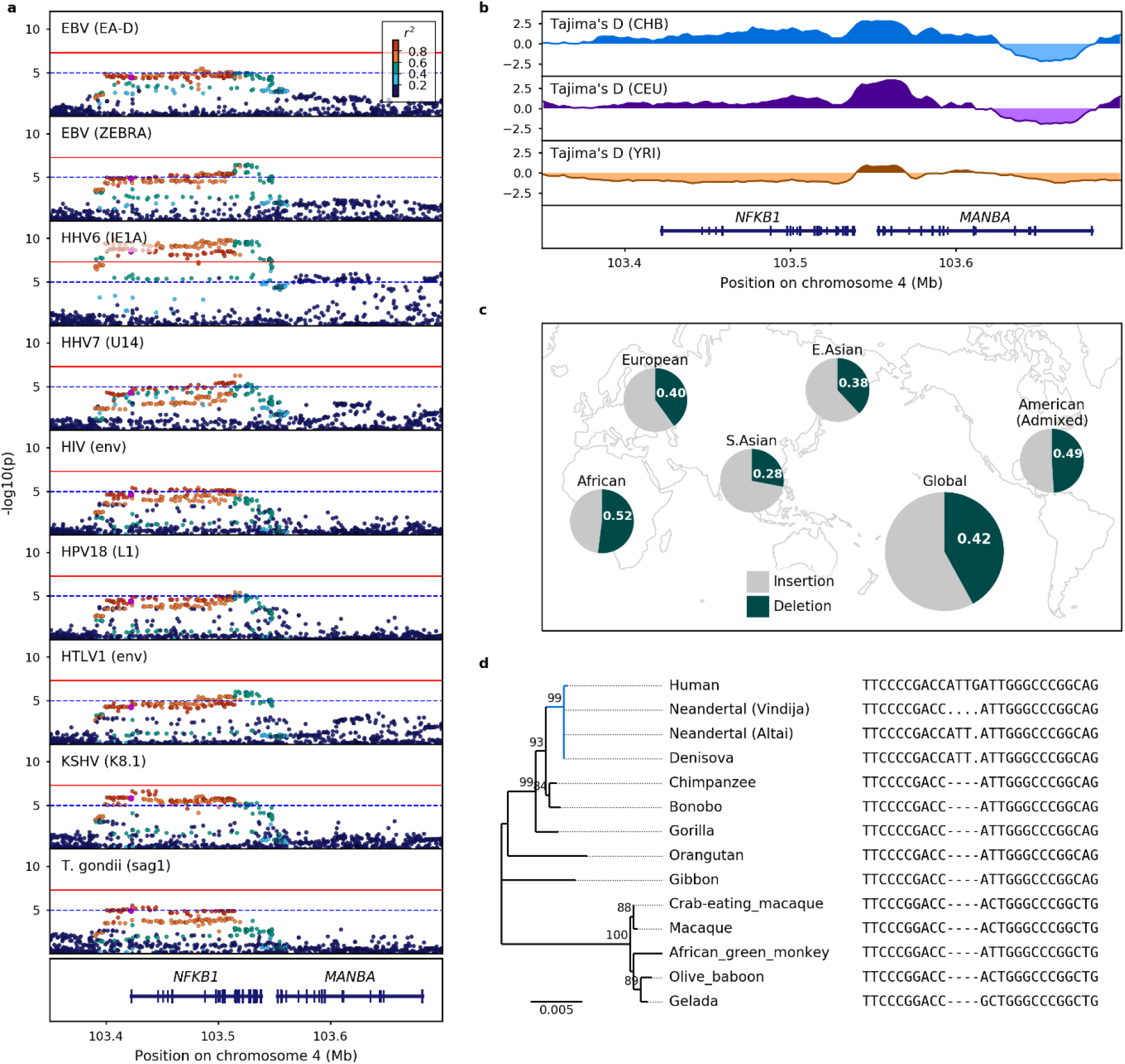
rs28362491 is an ancient variant affecting antibody responses to a range of pathogens. (a) Regional association plots showing the consistency of associations across the *NFKB1*/*MANBA* region on chromosome 4 in UK Biobank. Variants are coloured based on linkage with rs28362491 (indicated in purple). The upper red line indicates genome-wide significance (*P* = 5×10^−8^), and the lower blue dashed line indicates suggestive significance (*P* = 1×10^−5^). (b) Binned Tajima’s D selection statistics for 30 kb sliding windows across *NFKB1*/*MANBA* for 1000 Genomes Han Chinese in Beijing (CHB), Central European (CEU), and Yoruba in Ibadan, Nigeria (YRI) populations. A positive Tajima’s D is indicative of balancing selection, while a negative value suggests a recent selective sweep or population expansion. (c) Global population frequencies of rs28362491 based on allele frequencies from the 1000 Genomes project. (d) Phylogeny of *NFKB1* gene sequence across primates and hominids (highlighted in blue). The nucleotide sequence surrounding the rs28362491 indel variant is shown alongside. Dashes indicate the ancestral deletion variant; dots indicate potentially missing data from ancient DNA sequencing.

To identify the ancestral source of the variant, we compared the region immediately upstream of *NFKB1* across primates and early hominids and determined that the deletion allele is the ancestral variant (Fig. 2d), with the insertion arising early in hominid evolution between 550 ka and 8.4 million years ago. This was some time after the divergence of early hominids from chimpanzees (4.7–8.4 million years; ^28,29^), but before the divergence of archaic and modern humans (765–550 ka; ^30^). That this variant has been maintained throughout hominid evolution suggests that the insertion, and the resulting promoter binding region are evolutionarily significant, further supporting the presence of balancing selection acting on this locus.

### *NFKB1* is associated with dysregulation of inflammatory responses, balancing susceptibility to infections with risk of allergy

We found that the direction of effect for rs28362491 was consistent across 9 antigens for which we observed a signal for serum antibody responses in UK Biobank (EBV (EAD and ZEBRA), HHV-6 (IE1A), HHV-7 (U14), HPV-18 (L1), HTLV-1 (env), KSHV (K8.1), *T. gondii* (sag1)), with the deletion allele (rs28362491:delATTG) being associated with lower antibody responses in UK Biobank. Given this observation, with our observed replication in CoLaus/PsyCoLaus, and the published associations with multiple infectious or inflammatory conditions (Extended Data Table 3), we further tested for the effect of rs28362491:delATTG specifically on susceptibility to infection or inflammatory disease in UK Biobank, using prevalent and incident hospital inpatient (ICD-10) or self-reported cases of infection and inflammatory conditions in 487,297 participants (case definitions outlined in Supplementary Tables 3 and 4). Firstly, we replicated the previously published associations with diverse traits such as allergic rhinitis and Ulcerative colitis as reported in Extended Data Table 3 confirming the variant’s role in a variety of traits (Fig. 3A). Then we explored the effect of the variant on reported traits related to the measured antibody responses. For example we found that although rs28362491:delATTG was consistently associated with reduced antibody levels against all measured HPV antigens, there was an increased odds of reporting cervical intraepithelial neoplasia (OR = 1.17 (1.02-1.18)) or having an ICD code of papillomavirus (OR = 1.09 (1.02-1.34)) (Fig. 3B). We observed a similar pattern for EBV where the deletion was associated with reduced responses against multiple EBV antigens (e.g. EA-D: OR = 0.94 (0.92-0.97)), yet there was a trend towards increased odds of self-report of EBV infection/infectious mononucleosis (OR = 1.07 (0.98-1.18)) and multiple sclerosis (which has been plausibly causally linked with EBV infection; OR = 1.05 (0.98-1.11) in our UKB data), as well as a similar effect for *H. pylori*.

**Fig. 3:**
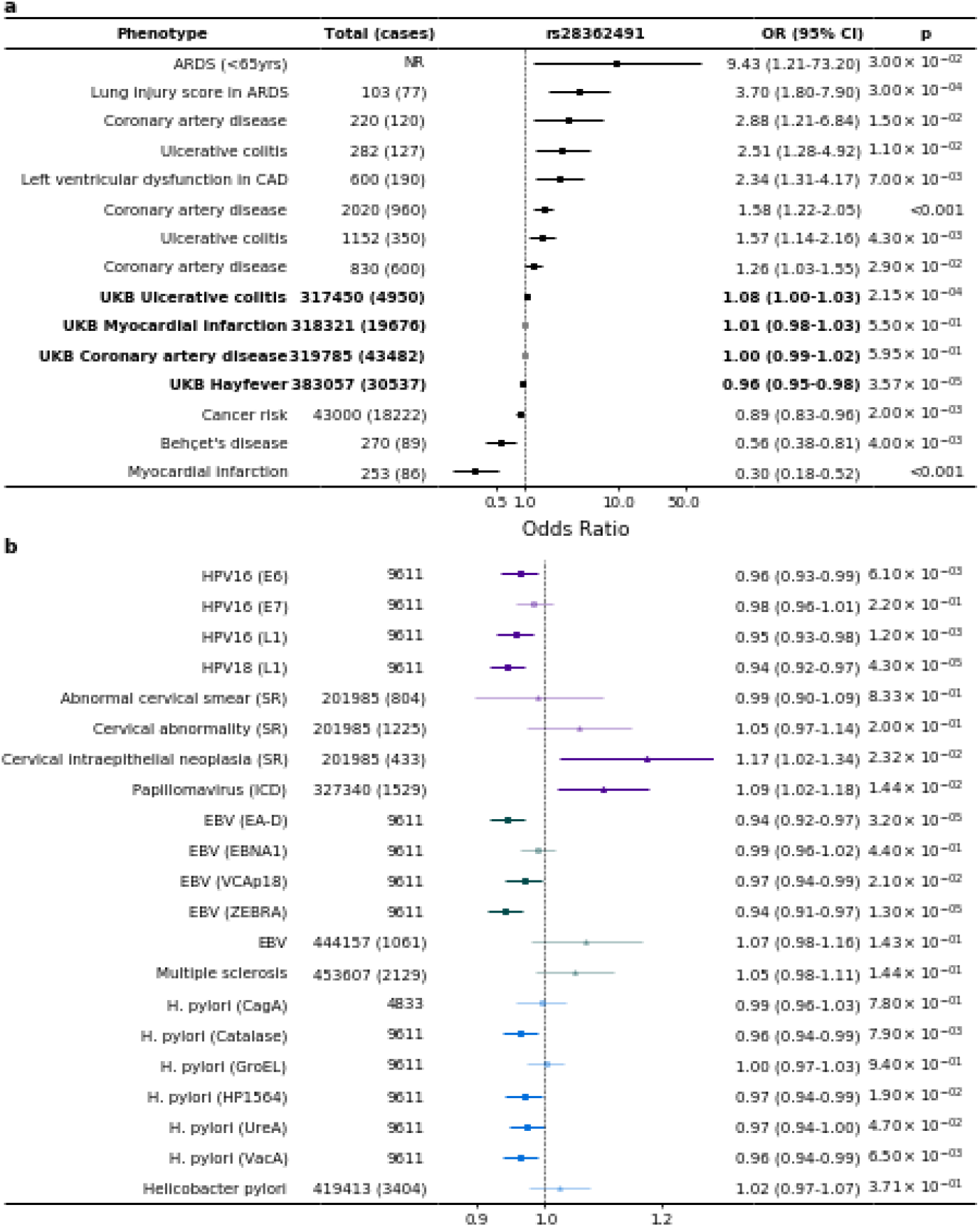
rs28362491:delATTG has a small, consistent effect, associated with increasing the risk of a range of infections and disease associated with dysregulation of inflammation, while protecting against exacerbated immune responses and allergy. (a) Published associations between rs28362491 and disease, and comparable summary stats for UKB (highlighted in bold). References listed in Extended Data Table 3. (b) Odds ratios and 95% confidence intervals for association between rs28362491, normalised antibody response against 14 antigens and 7 health-record derived disease traits. Square markers indicate quantitative antibody response phenotypes, and triangle markers indicate phenotypes derived from health record data (ICD-10 and self-report). Solid colours indicate traits with a *P* < 0.05. ARDS: Acute respiratory distress syndrome; CAD: Coronary artery disease; ICD: Phenotype is derived from ICD-10 health record data; SR: Phenotype is derived from self-reported health record data.

We then proceeded to test the association of the variant with a wider number of infectious and inflammatory diagnoses. Out of 39 infections tested, the deletion allele increased risk of infection in three, and decreased the risk of infection in one (Neisseria meningitidis (ICD), OR = 0.704 (0.509-0.974); *P* = 3.43×10^−2^). Since many infections were limited in number, we also tested the effect of the deletion allele against a phenotype consisting of any infection and found that overall there was a significant increase in risk associated with the deletion (OR = 1.014 (1.001-1.027), *P* = 3.31×10^−2;^ Supplementary Table 5). Furthermore, we found that the same variant was associated with an increased risk of diseases associated with dysregulation of inflammatory responses and chronic inflammation, such as psoriasis. In contrast, we found that the deletion allele was protective against allergic disease such as allergic rhinitis and asthma which are associated with acute inflammatory responses.

Altogether, these results demonstrate that this variant is associated with diverse groups of infectious or allergic diagnoses. It appears that the deletion tends to increase the risk of infectious or chronic inflammatory diagnoses, possibly associated with reduced antibody responses. Conversely, the insertion is associated with increased antibody response (and thus reduced risk of infectious diagnoses) but will come at an increased risk of allergic conditions. Given these observed divergent patterns of association with risk of inflammatory and infectious or allergic disease, alongside of our observation of balancing selection, we proceeded to test for risk of mortality in UK Biobank participants due to allergic, infectious or inflammatory disease. Although low in frequency given the relative immaturity of the UK Biobank cohort study, we observed similar trends of a protective effect of the deletion variant against death from allergic disease (OR = 0.846 (0.658-1.088), *P* = 0.192) and an increased risk of infection (OR = 1.065 (0.982-1.155), *P* = 0.129) and inflammation (OR = 1.038 (0.991-1.087), *P* = 0.117) (Supplementary Table 6).

### *NFKB1* might act as a master regulator of cellular transcription in immune cells

Having found significant evidence of the rs28362491:delATTG variant being associated with phenotypic disease endpoints and our intermediate antibody traits, we next sought to use these associations to understand the likely molecular pathways disrupted leading to increased infection risk. We first investigated the effect of rs28362491:delATTG on white blood cell counts and function (Supplementary Table 7). Using cell count data from UK Biobank, we observed that this variant was not significantly associated with lower overall white blood cell counts (*P* = 0.96), but instead had a higher total number of lymphocytes (beta = 1.98×10^−2^, *P* = 3.30×10^−24^) and eosinophils (beta = 6.49×10^−3^, *P* = 9.30×10^−4^) than those without. The deletion carriers also had lower counts of basophils (beta = -1.25×10^−2^, *P* = 2.00×10^−9^), monocytes (beta = -1.50×10^−2^, *P* = 2.70×10^−15^), neutrophils (beta = -7.27×10^−3^, *P* = 2.50×10^−4^), and platelets (beta = -5.79×10^−3^, *P* = 1.30×10^−3^) (Fig. 4).

**Fig. 4:**
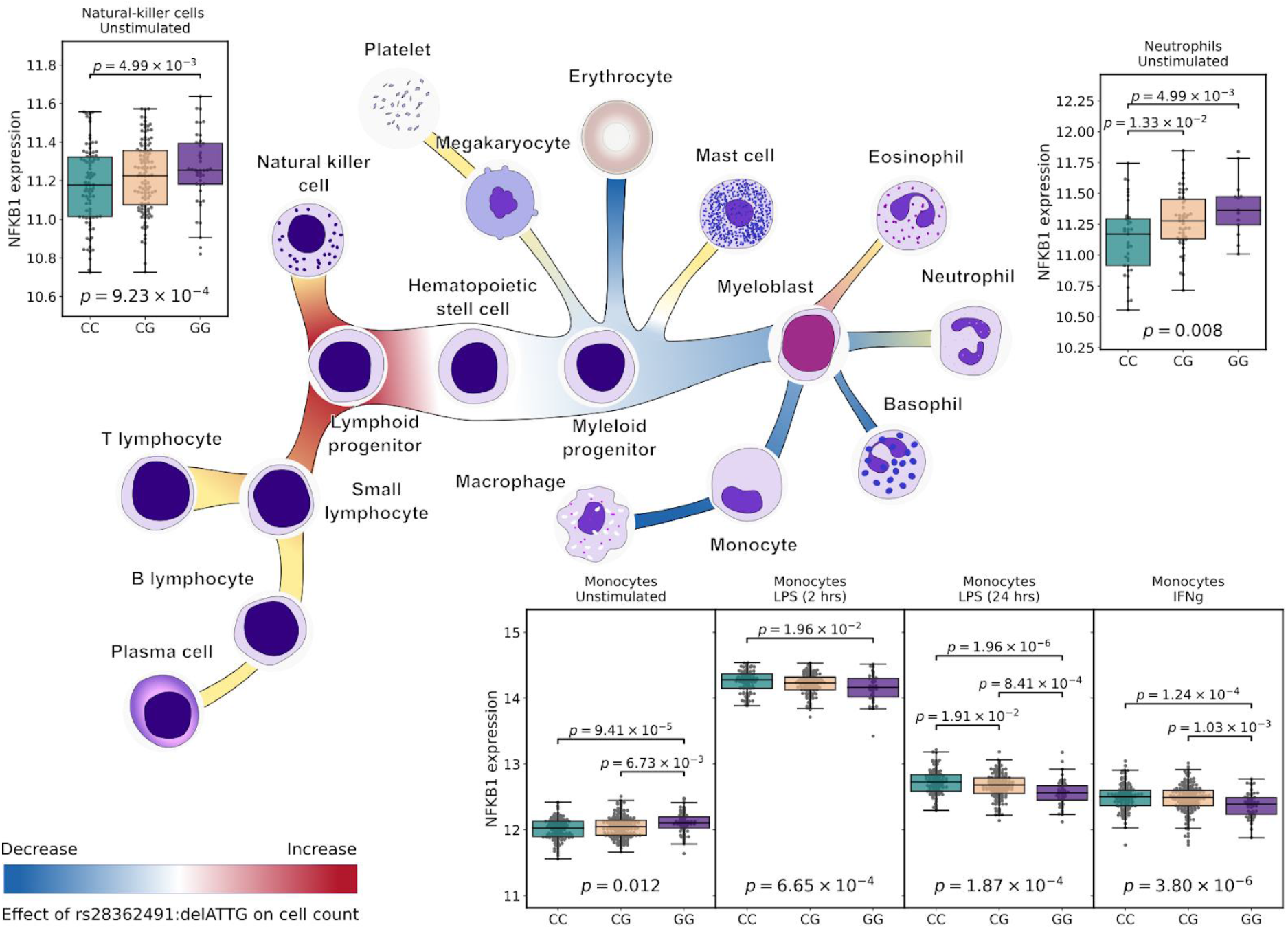
rs28362491 variation is also associated with differential haematopoiesis and *NFKB1* expression in white blood cells. Effect of rs28362491:delATTG on white blood cell counts is indicated by red-blue shading. Yellow shaded regions indicate a lack of information on cell counts (mast cells, B and T cells) or signals that did not reach genome-wide significance (NK cells, platelets, neutrophils, and eosinophils). Plot insets show differential expression by genotype at strongly linked 5‘UTR SNP rs72696119 (delATTG linked with G), eQTL p-values, and p-values for pairwise T-tests for differential expression by genotype. Clockwise from top-left: Unstimulated NK-cells, unstimulated neutrophils, unstimulated and stimulated monocytes. Expression for B and T cells not shown.

We next interrogated the differential expression of *NFKB1* in diverse blood cell types and found that the effect varied depending on the stimulation state of the cell. As rs28362491 is upstream of *NFKB1*, we used a tightly linked SNP (rs72696119, r^2^ = 0.999 D’ = 0.999) in the 5’ untranslated region (UTR) of *NFKB1* as a tagging SNP for eQTL analyses. We found that rs72696119 is a cis-eQTL for *NFKB1* in resting monocytes (*P* = 1.20×10^−2^), neutrophils (*P* = 8.00×10^−3^), NK cells (*P* = 9.23×10^−4^) and naive CD4+ and CD8+ T-cells (*P* = 3.50×10^−4^ and *P* = 1.40×10^−3^) but not in B-cells (*P* = 0.99). All significant loci in all tested analyses were found to colocalise.

### Summary

The genetic architecture underlying susceptibility to a host of infectious diseases is being resolved to greater detail. Loci including the MHC, immunoglobulin loci and FUT2 secretor status are becoming associated with a larger number of infectious traits when detected through multiple study formats, either through case-control analyses of clinical disease definition, self-report or antigen-specific diagnostics. Here we identify rs28362491 in the promoter region of *NFKB1*, which is common in all populations (global allele frequency (del) = 0.42), that incurs a subtle, yet demonstrable effect across both the UK Biobank population and, where available, other population datasets, modulating the risk of a range of infections. We found that this ancestral *NFKB1* variant predates speciation of early humans and has been maintained in human populations at a high frequency since with significant evidence of balancing selection occurring over long evolutionary time. As expected, given the known pleiotropy of NF-κB signalling, our study provides significant evidence of the delicate balance between immune response to infection and inflammatory processes, where disruptions can tip the balance towards exacerbated immune responses and allergy or towards increased risk of infection and impaired resolution of inflammation. There are likely multiple mechanisms underlying this effect, involving differential haematopoiesis and immunoglobulin class switching driven by modulation of *NFKB1* gene expression.

On the basis of our functional work, we suggest that reduction of *NFKB1* expression, driven by the deletion variant, favours development of lymphoid progenitor cells and lymphoid cell lineages over myeloid lineages during haematopoiesis. NF-κB activity regulates B-cell development and maturation and is important for T-cell co-stimulation ^31^. Despite moderate increases in the relative number of circulating lymphocytes, reduced *NFKB1* expression in T-cells may result in impaired T-cell function, thereby making them less responsive to infection. Variants tightly linked to rs28362491 have been associated with a reduced IgA/IgG:IgM ratio (Extended data Table 3), suggesting that reduced *NFKB1* expression may also affect B-cell class switching during activation ^13,20^, leading to the reduced IgG antibody levels observed in our study.

Interestingly, carriage of rs28362491:delATTG, was associated with higher levels of *NFKB1* gene expression in all cell types except B and T cells (Fig. 4 insets). This was unexpected given that the deletion allele results in the disruption of a promoter binding site. This effect was reversed in monocytes stimulated with interferon gamma (IFNγ) or lipopolysaccharide (LPS), where lower *NFKB1* expression was observed after stimulation. Although the mechanisms for this change in expression profile are unclear, it might also be related to *NFKB1* self-regulation, as with the dysregulation of inflammation, where disruption of *NFKB1* transcription disrupts NF-κB activity, resulting in altered expression profiles in individuals carrying the deletion allele. Our results support the role of *NFKB1* as a master regulator of cellular transcription that is finely balanced depending on cell state and stimulation.

The observed inverse effect in allergy compared with infection and inflammatory disease may be a result of impaired resolution of inflammation stemming from a dysregulation of *NFKB1* expression as a result of disrupted promoter binding. NF-κB family members and downstream gene products play a key role in induction and regulation of inflammatory responses, and as such, are tightly regulated, possibly by relative ratios of the NF-κB p50-p65 (*NFKB1* encodes p50, p65 is encoded by *RELA*) and p50-p50 dimers ^32^. Taken together with our cellular findings, it would appear that in the absence of haploinsufficiency, variation and regulation of expression is more important than overall *NFKB1* activity in altering risk of infection or inflammatory disease. However, follow-up studies will be required to fully dissect the effects of mild disruptions to NF-κB pathways particularly since many of our observations are based on cross-sectional analyses and therefore longitudinal analyses should be prioritised for the future.

Although we present evidence from a variety of sources and study formats in support of our observation, that strengthens the credibility of the association, our work does not prove definitively that, firstly, rs28362491:delATTG is the primary driving variant, nor that it is causal in all phenotypes studied. However, our fine mapping methods would make it the most likely candidate and historic transcription binding studies would make it the most plausible functional modulator of expression. Moreover, the subtle yet extensive pleiotropy observed with carriage of the alternate alleles of this variant makes contemporary methods of causal inference such as Mendelian Randomisation unreliable, and deeper validation of our hypothesis of mechanism highly challenging to dissect.

Identifying rs28362491:delATTG as a robust association with the multiple traits presented here has required data from a substantial number of individuals demonstrating that, although the effect of the variant at an individual scale may be subtle, the variation is very important on a population level influencing susceptibility to a range of diseases important to human health. These findings are likely to be important for future disease prediction scores and potentially from a therapeutics perspective building on our knowledge of the NFKB gene complex from primary immunodeficiency work where rare but large effect variants contribute to disease susceptibility. However, the NF-κB pathway will be difficult to target therapeutically since an altered balance will encourage either the development of infection or allergy. However, an acute disruption in favour of increased expression in cases of, for example adjuvantation during vaccination, could offer an opportunity for some infectious disease prevention on a public health scale.

## Methods

### Serology and GWAS

We used a Multiplex Serology platform described in Mentzer et. al. ^22^ to measure antibody responses against 45 antigens from 20 viral, bacterial, and protozoan pathogens present in the UK ^33^(Supplementary Table 1). Viral pathogens included on the panel included herpes simplex virus types 1 and 2 (HSV-1 and 2), varicella-zoster virus (VZV), Epstein-Barr virus (EBV), human cytomegalovirus (CMV), human herpesvirus 6 and 7 (HHV-6 and 7), Kaposi’s sarcoma-associated herpesvirus (KSHV), hepatitis B virus (HBV), hepatitis C Virus (HCV), human T-lymphotropic virus 1 (HTLV-1), human immunodeficiency virus 1 (HIV-1), BK and JC polyomaviruses (BKV and JCV), Merkel cell polyomavirus (MCV), and human papillomavirus 16 and 18 (HPV16 and 18). Other pathogens assayed were the bacterial pathogens *Chlamydia trachomatis* and *Helicobacter pylori*, and protozoan parasite *Toxoplasma gondii*.

For all association tests, we used the genotyped and imputed autosomal variant datasets available for the UK Biobank cohort ^34^. This consists of 805,426 high-quality genotyped variants, generated from the UK BiLEVE and UK Biobank Axiom arrays, and 93,095,623 imputed autosomal variants imputed using the Haplotype Reference Consortium (HRC), UK10K and 1000 Genomes phase 3 reference panels ^35–38^. For the serology phenotypes, we selected a subset of 9,611 random, unrelated individuals that had genetic data and serum samples available. Antibody responses across all participants regardless of serostatus were normalised using a rank-based inverse normal transformation. We measured the Spearman’s rank correlation between traits to determine whether there was any evidence of correlation between antibody traits (Supplementary Fig. 1).

Association analyses using imputed genotype data and both the untransformed and normalised data from quantitative serology traits were carried out using linear mixed models as implemented in BOLT-LMM (minor allele frequency (MAF) > 0.01, imputation infoscore > 0.3; ^39^) including age at recruitment and genetic sex as covariates. To identify the genomic regions and candidate genes associated with antibody response to each of the tested antigens, we first identified SNPs associated with tested antigens based on genome-wide significance (*P* < 5×10^−8^) and suggestive significance (*P* < 1×10^−5^). Starting with the most significant SNPs (peak SNP), we defined associated regions for each antigen based on calculated linkage disequilibrium from best-guess genotypes (R2 ≥ 0.2, calculated with PLINK; ^40^) between the peak SNP and other SNPs reaching suggestive significance for the dataset. This was repeated for each SNP that was not linked with a previous peak SNP. We then merged overlapping regions across all traits to define independent regions of association for further analysis.

### Finemapping of GWAS signals

For regions containing SNPs with an association p-value less than the Bonferroni corrected p value (*P* < 1.1×10^−9^), we then performed conditional analyses to determine if there were multiple independent signals present within the region. Analyses were carried out in GCTA using the --mlma option and adjusting for allelic dosage of the top SNP for each associated trait ^41^. These gene dosages were also used to test for interdependence between these associated regions and other regions of interest.

FINEMAP was used (default settings, R2 = 0.8; ^42^) to identify potential causal groups (Bayes factor ≥ 2) of SNPs within each of these regions based on summary statistics for each of the associated traits. These were then compared across antigens and merged to define haplotypes of SNPs based on shared SNPs and direction of effect. Causal groups sharing SNPs were only merged if the direction of effect across all antigens were in a consistent direction relative to the minor allele.

Potential causal SNPs were identified from haplotypes shared across multiple antigens. To do this, we first checked the location of SNPs relative to the Ensembl GRCh37.p13 gene annotations to identify the nearest genes. The predicted effects of each variant were then retrieved using the Ensembl Variant Effect Predictor^43^.

### Cohorte Lausannoise replication study and meta-analysis

The Cohorte Lausannoise (CoLaus/PsyCoLaus) study includes 6,188 individuals of European ancestry living in Lausanne, Switzerland who were randomly selected from the general population. Recruitment and sampling protocols are described by Firmann et. al. ^44^. Genotyping data for 5,399 individuals were obtained using the Affymetrix Axiom Biobank array. Following quality control steps to remove poorly performing variants and samples, imputed genotypes were generated using the Sanger Imputation Service ^38^. Genotypes were phased using EAGLE2 (v2.0.5; ^37^) and the HRC panel ^38^, and imputation was carried out using the HRC reference panel, 1000 Genomes Phase 3 data, and the UK10K reference panel ^35,36^. Serology phenotypes were generated using a similar Multiplex Serology Panel to the UK Biobank data as described above ^16,33^.

Genome-wide association analyses were carried out using 4216 individuals with imputed genotype data and serology data, and linear mixed models as implemented in GCTA (1.91.3beta; ^41^) and thresholded high quality imputed genotype data (imputation infoscore > 0.8, MAF > 0.01, *P*_HWE_ < 10×10^−7^). Age, sex, and the top three principal components were included as covariates. For antigens with a signal at *NFKB1* and that were also available from the CoLaus/PsyCoLaus cohort, we performed GWAS that were common across the two cohorts, we extracted the set of intersecting SNPs for meta-analysis using Metasoft (v2.0.1, ^45^).

### Infection and inflammation associations in UK Biobank

We used self-reported non-cancer illness (Field ID: 20002) and prevalent and incident hospital inpatient ICD-10 codes (Field ID: 41270) to define cases and controls for further infection and inflammation related phenotypes within UK Biobank. For self-reported conditions, cases and controls were drawn from participants who reported the presence or absence of non-cancer related conditions, and had genetic data available, excluding those who did not respond to the question. Controls for infection-related traits derived from self-reported data were defined as individuals who had reported no major infections for infection-related phenotypes with genetic data available. Controls for other immune-related traits were drawn from all unaffected individuals with genetic data.

Similarly, potential cases and controls based on ICD-10 data were drawn from those individuals with hospital inpatient diagnoses and genetic data available. Case definitions are outlined in Supplementary Tables 3 and 4. Controls for ICD-10 derived phenotypes were drawn from individuals with hospital inpatient and genetic data, who were not affected by the trait of interest. In the case of infection-related traits, the selection was further restricted to those who had not reported a major infection, or with no ICD codes relating to infection (e.g. Chapter I (A00-B99), Chapter X: Diseases of the respiratory system: Acute upper respiratory infections (J00-J06), Influenza and pneumonia (J09-J18), Other acute lower respiratory infections (J20-J22)).

Where equivalent phenotypes were present in both self-reported and ICD-10 data, a joint phenotype was defined using all possible cases and controls. Similarly, summary phenotypes for allergy, infection, and inflammation were defined using all available cases and controls. Due to the small numbers of cases across many phenotypes, we used only those with more than 50 cases.

Association tests for these traits were carried out using linear mixed models as implemented in SAIGE (v 0.36.3.2; ^46^), also including age at recruitment and genetic sex as covariates, except for traits where only one sex was present in the cases, in which case sex was not included, but controls were drawn only from individuals of that sex (see Supplementary Tables 3 and 4 for more information). Due to relatively small numbers of individuals for which cause of death was available (approximately 25,000), we used logistic regression to assess the relationship between rs28362491:delATTG and the contribution of infection or inflammatory conditions to cause of death. Similar to the GWAS analysis described above, we used imputed dosages of rs28362491:delATTG, with age and sex as covariates. We also restricted analysis to individuals in the white British subset as defined by UK Biobank.

### Association with blood cell counts

Association tests for blood cell counts were carried out on the full UK Biobank cohort using transformed cell count data and imputed genotypes. Traits included red blood cell (RBC) count, white blood cell (WBC) count, platelet, basophil, eosinophil, lymphocyte, monocyte, and neutrophil counts. Values were normalised using a rank-based inverse normal transformation prior to analysis. Association tests were carried out using BOLT-LMM including age at recruitment and genetic sex as covariates.

### eQTL analysis

To assess the regulatory function of rs28362491, we used the tightly linked, 5‘UTR SNP rs72696119. We correlated rs72696119 genotype with *NFKB1* RNA expression in naïve and stimulated primary immune cell subsets from healthy European adults, using data from published and unpublished expression quantitative trait locus (eQTL) studies. CD56+CD3-NK cells, CD19+ B cells, CD14+ monocytes, CD16+ neutrophils were separated, genotyped, and gene expression quantified as previously described ^47–50^. Following quality control and normalisation of gene expression data, we correlated rs72696119 genotype with *NFKB1* RNA expression in each cell type: B-cells (n = 279), NK cells (n = 245), neutrophils (n = 101), naïve monocytes (n = 414), and stimulated monocytes (LPS 2 hours, n = 261; LPS 24 hours, n = 322; IFNγ 24 hours, n = 367). *NFKB1* expression was correlated with genotype by linear regression and analysis of variance (ANOVA), including the first 25 principal components of gene expression data in each cell-type/condition to account for confounding variation. P-values are calculated with F-tests (1 d.f.). Statistical analysis was performed in R. We used publicly available eQTL data from the DICE database for CD4+ and CD8+ T cells to examine association of rs72696119 with *NFKB1* expression in naïve T-cells (^51^; https://dice-database.org/).

### Colocalisation of eQTL and GWAS signals

Colocalisation testing between eQTL signals and antibody and blood cell GWAS association signals was carried out using coloc (v5.0.1,^52^) with susieR (v0.11.92, ^53,54^) and the intersection of variants from all GWAS phenotypes and the eQTL traits from the region immediately upstream of and across NFKB1. LD was calculated independently for each dataset using PLINK. Colocalisation was tested between pairs of antibody traits, between HHV6 (IE1A) and blood cell traits, between HHV6 (IE1A) and eQTL datasets, and between related blood cell traits and eQTL datasets (e.g. monocyte count and unstimulated and stimulated monocyte eQTL datasets) (Supplementary table 8).

### *NFKB1* phylogenetics

We extracted the first exon of *NFKB1* and 500bp immediately upstream from 11 high quality primate genomes present in Ensembl. These sequences were aligned using ClustalW ^55^ and manually inspected to determine the presence of the 4bp indel variant. To confirm the absence of the insertion in great apes, we used variation data for six great ape species: Eastern lowland gorilla (*Gorilla beringei*; n = 3), Western lowland gorilla (*Gorilla gorilla*; n = 3), Bonobo (*Pan paniscus*, n = 13), Chimpanzee (*Pan troglodytes*; n = 14), Sumatran orangutan (*Pongo abelii*; n = 5), and Bornean orangutan (*Pongo pygmaeus*; n = 5) ^56^. We also used variation data from three Neanderthals, one Denisovan genome, and three ancient humans ^30^ to estimate the latest common ancestor where the indel variant may have arisen.

### Selection testing

Tajima’s D population genetic summary statistics and rank scores for the CEU, YRI, and CHB populations were retrieved from the Human Positive Selection Browser ^26^ for *NFKB1* and regions 200kb upstream and downstream of the gene.

## Data Availability

Summary statistics are available to download from Zenodo.

https://doi.org/10.5281/zenodo.7347714

https://doi.org/10.5281/zenodo.7347792

## Data availability

GWAS summary statistics are available on Zenodo. Serology data:

https://doi.org/10.5281/zenodo.7347714; Heath record and Blood cell count data:

https://doi.org/10.5281/zenodo.7347792

## Acknowledgements

We thank the UK Biobank participants and all individuals involved in recruitment, data and sample collection, data curation and release. This research has been conducted using the UK Biobank Resource under Application Number 43920. This research has been conducted using data from the CoLaus/PsyCoLaus Study.

AJM was supported by a Wellcome Trust Fellowship with reference 106289/Z/14/Z and the National Institute for Health Research (NIHR) Oxford Biomedical Research Centre (BRC). AYC was supported by the Research Councils UK Newton Fund Award with reference MR/N028937/1. AJ-K and AM-E were supported by The Mexican Biobank Project (CONACYT-Newton Fund grant number FONCICYT/50/2016). JJG is funded by a National Institute for Health Research (NIHR) Clinical Lectureship. JF was supported by the Swiss National Science Foundation, grant #175603. The research was supported by the Wellcome Trust Core Award Grant Number 203141/Z/16/Z with additional support from the NIHR Oxford BRC. The views expressed are those of the authors and not necessarily those of the NHS, the NIHR or the Department of Health.

## Author contributions

AYC and AJK carried out the GWAS of antibody traits; AYC performed additional analysis of *NFKB1*-related traits. NB, MH, TJL, TW, and AJM performed the antibody data generation and NB, TJL, TW, and AJM curated the data. AC, MH, TJL contributed to the methods development. JJG, BPF, and JCK carried out analysis of the eQTL data. FH and JF carried out analysis of the CoLaus/PsyCoLaus cohort. TW, AVSH, and AJM designed the experiments. GM, AME, TW, AVSH, and AJM supervised the research and provided funding.

## References

1. COVID-19 Host Genetics Initiative. Mapping the human genetic architecture of COVID-19. Nature 2021 1–8 (2021).

2. The Severe Covid-19 GWAS Group. Genomewide Association Study of Severe Covid-19 with Respiratory Failure. NEJM 383, 1522–1534 (2020).

3. Pairo-Castineira, E. et al. Genetic mechanisms of critical illness in COVID-19. Nature 2020 591:7848 591, 92–98 (2020).

4. Davila, S. et al. Genome-wide association study identifies variants in the CFH region associated with host susceptibility to meningococcal disease. Nature Genetics 42, 772–776 (2010).

5. Duggal, P. et al. Genome-Wide Association Study of Spontaneous Resolution of Hepatitis C Virus Infection: Data From Multiple Cohorts. Annals of Internal Medicine 158, 235–245 (2013).

6. Dunstan, S. J. et al. Variation at HLA-DRB1 is associated with resistance to enteric fever. Nature Genetics 46, 1333–1336 (2014).

7. Gilchrist, J. J. et al. Risk of nontyphoidal Salmonella bacteraemia in African children is modified by STAT4. Nature Communications 9, (2018).

8. Vergara, C. et al. Multi-Ancestry Genome-Wide Association Study of Spontaneous Clearance of Hepatitis C Virus. Gastroenterology 156, 1496-1507.e7 (2019).

9. Wang, Z. et al. A large-scale genome-wide association and meta-analysis identified four novel susceptibility loci for leprosy. Nature Communications 7, 13760 (2016).

10. Zhang, F. et al. Identification of two new loci at IL23R and RAB32 that influence susceptibility to leprosy. Nature Genetics 43, 1247–1251 (2011).

11. Tian, C. et al. Genome-wide association and HLA region fine-mapping studies identify susceptibility loci for multiple common infections. Nature Communications 8, 599 (2017).

12. VanBlargan, L. A., Goo, L. & Pierson, T. C. Deconstructing the Antiviral Neutralizing-Antibody Response: Implications for Vaccine Development and Immunity. Microbiology and Molecular Biology Reviews 80, 989–1010 (2016).

13. Jonsson, S. et al. Identification of sequence variants influencing immunoglobulin levels. Nat Genet 49, 1182–1191 (2017).

14. Png, E. et al. A genome-wide association study of hepatitis B vaccine response in an Indonesian population reveals multiple independent risk variants in the HLA region. Human Molecular Genetics 20, 3893–3898 (2011).

15. Hammer, C. et al. Amino Acid Variation in HLA Class II Proteins Is a Major Determinant of Humoral Response to Common Viruses. Am J Hum Genet 97, 738–743 (2015).

16. Hodel, F. et al. Human genomics of the humoral immune response against polyomaviruses. Virus Evolution 7, 1–11 (2021).

17. Kachuri, L. et al. The landscape of host genetic factors involved in immune response to common viral infections. Genome Medicine 12, 1–18 (2020).

18. Hayden, M. S. & Ghosh, S. NF-kappaB in immunobiology. Cell Res 21, 223–244 (2011).

19. Hayden, M. S., West, A. P. & Ghosh, S. NF-kappaB and the immune response. Oncogene 25, 6758–6780 (2006).

20. Tuijnenburg, P. et al. Loss-of-function nuclear factor kappaB subunit 1 (NFKB1) variants are the most common monogenic cause of common variable immunodeficiency in Europeans. J Allergy Clin Immunol 142, 1285–1296 (2018).

21. Butler-Laporte, G. et al. Genetic Determinants of Antibody-Mediated Immune Responses to Infectious Diseases Agents: A Genome-Wide and HLA Association Study. Open Forum Infect Dis 7, (2020).

22. Mentzer, A. J. et al. Identification of host–pathogen-disease relationships using a scalable multiplex serology platform in UK Biobank. Nature Communications 13, (2022).

23. Scepanovic, P. et al. Human genetic variants and age are the strongest predictors of humoral immune responses to common pathogens and vaccines. Genome Med 10, 59 (2018).

24. Jenner, R. G. & Young, R. A. Insights into host responses against pathogens from transcriptional profiling. Nat Rev Microbiol 3, 281–294 (2005).

25. Karban, A. S. et al. Functional annotation of a novel NFKB1 promoter polymorphism that increases risk for ulcerative colitis. Hum Mol Genet 13, 35–45 (2004).

26. Pybus, M. et al. 1000 Genomes Selection Browser 1.0: a genome browser dedicated to signatures of natural selection in modern humans. Nucleic Acids Res 42, D903–9 (2014).

27. Tajima, F. Statistical method for testing the neutral mutation hypothesis by DNA polymorphism. Genetics 123, 585–595 (1989).

28. Kumar, S., Filipski, A., Swarna, V., Walker, A. & Hedges, S. B. Placing confidence limits on the molecular age of the human-chimpanzee divergence. Proc Natl Acad Sci U S A 102, 18842–18847 (2005).

29. Patterson, N., Richter, D. J., Gnerre, S., Lander, E. S. & Reich, D. Genetic evidence for complex speciation of humans and chimpanzees. Nature 441, 1103–1108 (2006).

30. Prufer, K. et al. A high-coverage Neandertal genome from Vindija Cave in Croatia. Science (1979) 358, 655–658 (2017).

31. Kaileh, M. & Sen, R. NF-kappaB function in B lymphocytes. Immunol Rev 246, 254–271 (2012).

32. Mishra, A., Srivastava, A., Mittal, T., Garg, N. & Mittal, B. Role of inflammatory gene polymorphisms in left ventricular dysfunction (LVD) susceptibility in coronary artery disease (CAD) patients. Cytokine 61, 856–861 (2013).

33. Waterboer, T. et al. Multiplex Human Papillomavirus Serology Based on In Situ–Purified Glutathione S-Transferase Fusion Proteins. Clinical Chemistry 51, 1845–1853 (2005).

34. Bycroft, C. et al. The UK Biobank resource with deep phenotyping and genomic data. Nature 562, 203–209 (2018).

35. 1000 Genomes Project Consortium et al. A global reference for human genetic variation. Nature 526, 68–74 (2015).

36. Huang, J. et al. Improved imputation of low-frequency and rare variants using the UK10K haplotype reference panel. Nature Communications 6, 8111 (2015).

37. Loh, P. R. et al. Reference-based phasing using the Haplotype Reference Consortium panel. Nature Genetics 48, 1443–1448 (2016).

38. McCarthy, S. et al. A reference panel of 64,976 haplotypes for genotype imputation. Nature Genetics 48, 1279–1283 (2016).

39. Loh, P. R. et al. Efficient Bayesian mixed-model analysis increases association power in large cohorts. Nature Genetics 47, 284–290 (2015).

40. Chang, C. C. et al. Second-generation PLINK: Rising to the challenge of larger and richer datasets. Gigascience 4, 7 (2015).

41. Yang, J., Lee, S. H., Goddard, M. E. & Visscher, P. M. GCTA: a tool for genome-wide complex trait analysis. Am J Hum Genet 88, 76–82 (2011).

42. Benner, C. et al. FINEMAP: efficient variable selection using summary data from genome-wide association studies. Bioinformatics 32, 1493–1501 (2016).

43. McLaren, W. et al. The Ensembl Variant Effect Predictor. Genome Biology 17, 1–14 (2016).

44. Firmann, M. et al. The CoLaus study: a population-based study to investigate the epidemiology and genetic determinants of cardiovascular risk factors and metabolic syndrome. BMC Cardiovasc Disord 8, 6 (2008).

45. Han, B. & Eskin, E. Random-effects model aimed at discovering associations in meta-analysis of genome-wide association studies. Am J Hum Genet 88, 586–598 (2011).

46. Zhou, W. et al. Efficiently controlling for case-control imbalance and sample relatedness in large-scale genetic association studies. Nature Genetics 50, 1335–1341 (2018).

47. Naranbhai, V. et al. Genomic modulators of gene expression in human neutrophils. Nature Communications 6, 7545 (2015).

48. Fairfax, B. P. et al. Genetics of gene expression in primary immune cells identifies cell type-specific master regulators and roles of HLA alleles. Nature Genetics 44, 502–510 (2012).

49. Fairfax, B. P. et al. Innate immune activity conditions the effect of regulatory variants upon monocyte gene expression. Science (1979) 343, 1246949 (2014).

50. Gilchrist, J. J. et al. Natural Killer cells demonstrate distinct eQTL and transcriptome-wide disease associations, highlighting their role in autoimmunity. bioRxiv (2021) doi:10.1101/2021.05.10.443088.

51. Schmiedel, B. J. et al. Impact of Genetic Polymorphisms on Human Immune Cell Gene Expression. Cell 175, 1701-1715.e16 (2018).

52. Wallace, C. A more accurate method for colocalisation analysis allowing for multiple causal variants. PLOS Genetics 17, e1009440 (2021).

53. Wang, G., Sarkar, A., Carbonetto, P. & Stephens, M. A simple new approach to variable selection in regression, with application to genetic fine mapping. Journal of the Royal Statistical Society: Series B (Statistical Methodology) 82, 1273–1300 (2020).

54. Zou, Y., Carbonetto, P., Wang, G. & Stephens, M. Fine-mapping from summary data with the “Sum of Single Effects” model. bioRxiv (2021) doi:10.1101/2021.11.03.467167.

55. Thompson, J. D., Higgins, D. G. & Gibson, T. J. CLUSTAL W: improving the sensitivity of progressive multiple sequence alignment through sequence weighting, position-specific gap penalties and weight matrix choice. Nucleic Acids Res 22, 4673–4680 (1994).

56. Prado-Martinez, J. et al. Great ape genetic diversity and population history. Nature 499, 471–475 (2013).

57. Waage, J. et al. Genome-wide association and HLA fine-mapping studies identify risk loci and genetic pathways underlying allergic rhinitis. Nat Genet 50, 1072–1080 (2018).

58. Johansson, A., Rask-Andersen, M., Karlsson, T. & Ek, W. E. Genome-wide association analysis of 350 000 Caucasians from the UK Biobank identifies novel loci for asthma, hay fever and eczema. Hum Mol Genet 28, 4022–4041 (2019).

59. Bajwa, E. K. et al. An NFKB1 promoter insertion/deletion polymorphism influences risk and outcome in acute respiratory distress syndrome among Caucasians. PLoS One 6, e19469 (2011).

60. Ellinghaus, D. et al. Analysis of five chronic inflammatory diseases identifies 27 new associations and highlights disease-specific patterns at shared loci. Nat Genet 48, 510–518 (2016).

61. Yenmis, G. et al. Association of NFKB1 and NFKBIA polymorphisms in relation to susceptibility of Behcet’s disease. Scand J Immunol 81, 81–86 (2015).

62. Seidi, A., Mirzaahmadi, S., Mahmoodi, K. & Soleiman-Soltanpour, M. The association between NFKB1 - 94ATTG ins/del and NFKB1A 826C/T genetic variations and coronary artery disease risk. Mol Biol Res Commun 7, 17–24 (2018).

63. Lai, H.-M. et al. Genetic Variation in NFKB1 and NFKBIA and Susceptibility to Coronary Artery Disease in a Chinese Uygur Population. PLOS ONE 10, e0129144 (2015).

64. Adamzik, M. et al. Insertion/deletion polymorphism in the promoter of NFKB1 influences severity but not mortality of acute respiratory distress syndrome. Intensive Care Med 33, 1199–1203 (2007).

65. Dudding, T. et al. Genome wide analysis for mouth ulcers identifies associations at immune regulatory loci. Nat Commun 10, 1052 (2019).

66. Boccardi, V. et al. -94 ins/del ATTG NFKB1 gene variant is associated with lower susceptibility to myocardial infarction. Nutr Metab Cardiovasc Dis 21, 679–684 (2011).

67. Kawashima, M. et al. Genome-wide association studies identify PRKCB as a novel genetic susceptibility locus for primary biliary cholangitis in the Japanese population. Hum Mol Genet 26, 650–659 (2017).

68. Gonzalez-Serna, D. et al. Analysis of the genetic component of systemic sclerosis in Iranian and Turkish populations through a genome-wide association study. Rheumatology (Oxford) 58, 289–298 (2019).

69. Liou, Y. J. et al. Genome-wide association study of treatment refractory schizophrenia in Han Chinese. PLoS One 7, e33598 (2012).

70. Jostins, L. et al. Host-microbe interactions have shaped the genetic architecture of inflammatory bowel disease. Nature 491, 119–124 (2012).

71. Borm, M. E., van Bodegraven, A. A., Mulder, C. J., Kraal, G. & Bouma, G. A NFKB1 promoter polymorphism is involved in susceptibility to ulcerative colitis. Int J Immunogenet 32, 401–405 (2005).

72. Wang, D. et al. Genetic association between NFKB1 -94 ins/del ATTG Promoter Polymorphism and cancer risk: a meta-analysis of 42 case-control studies. Sci Rep 6, 30220 (2016).

73. Kanai, M. et al. Genetic analysis of quantitative traits in the Japanese population links cell types to complex human diseases. Nature Genetics 50, 390–400 (2018).

