## Supplementary data for "A frequent ancestral NFKB1 variant predicts risk of infection or allergy"

### Supplementary Figures

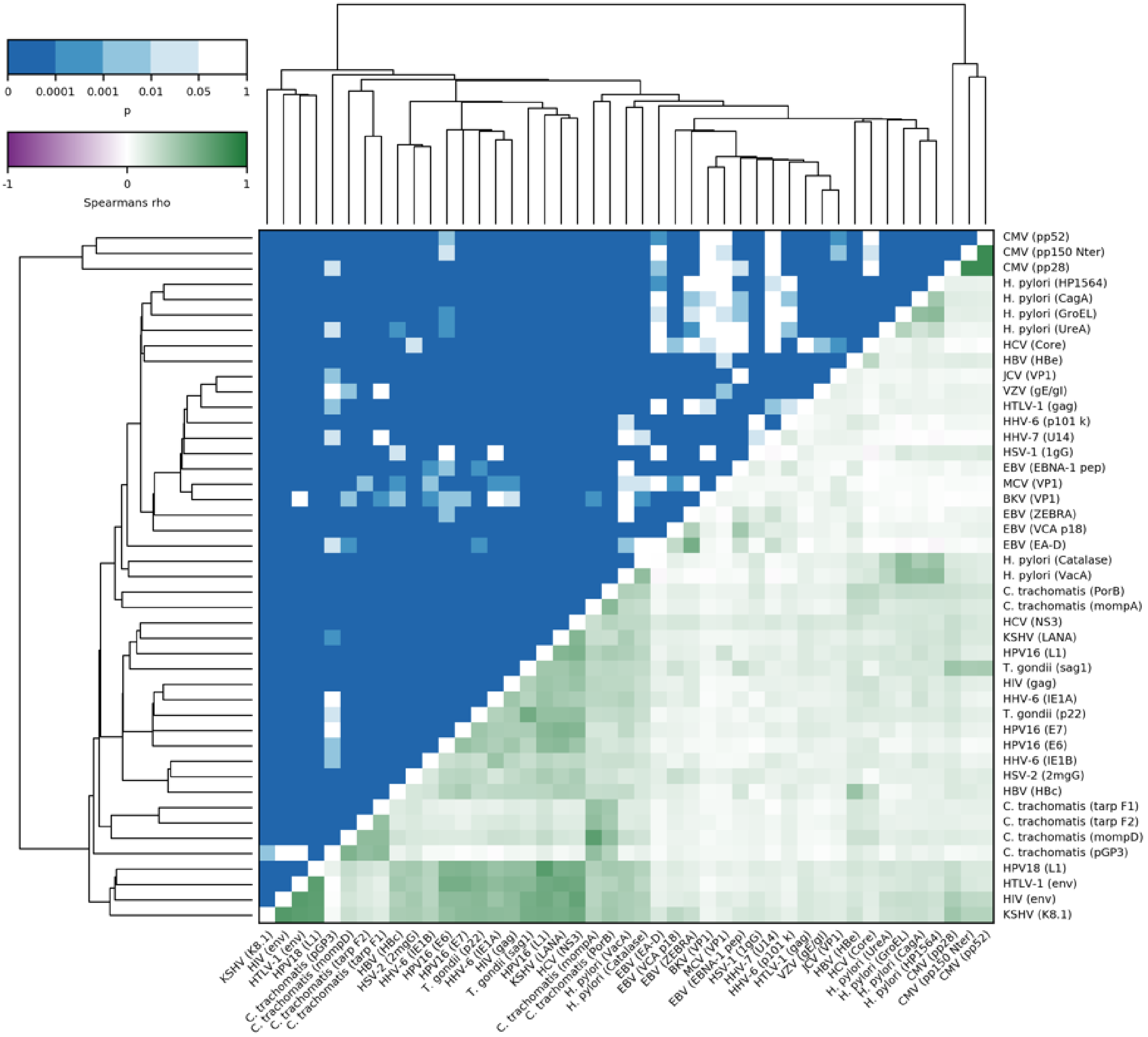


**Supplementary Fig. 1: All antibody responses were generally positively correlated.** Antibody responses to different antigens from the same pathogen tended to cluster together and were more strongly correlated with each other. No evidence of unexpected correlation between antibody traits was detected. Pairwise correlations between antibody responses are shown in the bottom triangle, p-values are indicated in the upper triangle

### Supplementary Tables

**Supplementary Table 1:** Details of the 45 antigens and 20 pathogens covered in the UK Biobank serology panel

| Infectious agent | Antigen | Likely Function | Reference |
|---|---|---|---|
| HSV-1 | 1gG | membrane glycoprotein | 1 |
| HSV-2 | 2mgGunique | membrane glycoprotein | 1 |
| VZV | gE/gI | envelope glycoproteins | 1 |
| EBV | VCAp18 | capsid protein | 1 |
| | EBNA1 | replication, latent viral infection | |
| | ZEBRA | replication activator | |
| | EA-D | replication (polymerase accessory subunit) | |
| CMV | pp150 (N-terminus) | tegument protein | 1 |
| | pp52 | DNA binding protein | |
| | pp28 | capsid protein | |
| HHV-6 | IE1B | potential transactivator | Validation ongoing |
| | IE1A | potential transactivator | |
| | p101k | potential tegument protein | |
| HHV-7 | U14 | potential tegument protein | Validation ongoing |
| KSHV | LANA | replication and long-term persistence | Validation ongoing |
| | K8.1 | structural glycoprotein | |
| HBV | HBc | core antigen | 2 |
| | HBe | soluble nucleocapsid associated antigen | |
| HCV | Core | structural antigen | 2,3 |
| | NS3 | protease and RNA helicase activity | |
| *Toxoplasma gondii* | p22 | surface protein | 2 |
| | sag1 | surface protein | |
| HTLV-1 | Gag | structural antigen | 2 |
| | Env | structural antigen | |
| HIV-1 | Gag | structural antigen | 4 |
| | Env | structural antigen | |
| BKV | VP1 | major capsid protein | 5–7 |
| JCV | VP1 | major capsid protein | 5–7 |
| MCV | VP1 | major capsid protein | 6,7 |
| HPV16 | L1 | major capsid protein | 8,9 |
| | E6 | oncogene | |
| | E7 | oncogene | |
| HPV18 | L1 | major capsid protein | 9 |

| | | | |
|---|---|---|---|
| *Chlamydia trachomatis* | mompD | structural antigen | 10,11 |
| | mompA | structural antigen | |
| | TarpDF1 | regulation of actin recruitment | |
| | TarpDF2 | | |
| | PorB | porin | |
| | pGP3 | virulence factor | |
| *Helicobacter pylori* | CagA (N-terminus) | pathogenesis | 12 |
| | VacA (C-terminus) | pathogenesis | |
| | HP1564 | cell envelope | |
| | GroEL | chaperonin | |
| | Catalase | detoxification | |
| | UreA | urease alpha subunit | |
| - | GST | used for background subtraction | 8 |

**Supplementary Table 2:** Cohort characteristics for UK Biobank and CoLaus/PsyCoLaus serology data

| Characteristic | UK Biobank | (%) | CoLaus/PsyCoLaus | (%) |
|---|---|---|---|---|
| **Age at recruitment** | | | | |
| 30-39 | 0 | 0.00 | 476 | 11.29 |
| 40-49 | 2250 | 23.41 | 1280 | 30.36 |
| 50-59 | 3145 | 32.72 | 1159 | 27.49 |
| 60-69 | 4158 | 43.26 | 991 | 23.51 |
| 70+ | 58 | 0.60 | 310 | 7.35 |
| **Sex** | | | | |
| Male | 4236 | 44.07 | 1946 | 46.16 |
| Female | 5375 | 55.93 | 2270 | 53.84 |
| **Reported ethnicity** | | | | |
| White | 9061 | 94.28 | 4216 | 100 |
| Asian | 234 | 2.43 | | |
| Black | 139 | 1.45 | | |
| Other | 134 | 1.39 | | |
| Not reported/missing | 43 | 0.45 | | |

**Supplementary Table 3:** Phenotype definitions of ICD-10 derived phenotypes

| Phenotype group | Phenotype | Cases | Controls | UK Biobank code(s) |
|---|---|---|---|---|
| Allergy | Allergic dermatitis | 129 | 397549 | L230, L231, L232, L234, L235, L236, L237, L238, L239 |
| Allergy | Asthma | 36040 | 361638 | J450, J451, J458, J459 |
| Allergy | Atopic dermatitis | 96 | 397582 | L200, L208, L209 |
| Allergy | Hayfever | 1784 | 325811 | J301, J302, J303, J304 |
| Allergy | Seborrhoeic dermatitis | 84 | 397594 | L210, L211, L218, L219 |
| Infection | *Aspergillus spp.* | 230 | 325811 | B440, B441, B442, B447, B448, B449 |
| Infection | *Bordetella pertussis* | 2 | 325811 | A370 |
| Infection | *Campylobacter spp.* | 527 | 325811 | A045 |
| Infection | *Candida spp.* | 3519 | 325811 | B370, B371, B372, B373, B374, B375, B376, B377, B378, B379 |
| Infection | *Clostridium difficile* | 963 | 325811 | A047 |
| Infection | *Corynebacterium diphtheriae* | 11 | 325811 | A360, A361, A362, A363, A368, A369 |
| Infection | CMV | 186 | 325811 | B271, B250, B251, B252, B258, B259 |
| Infection | EBV | 29 | 325811 | B270 |
| Infection | *Enterobius vermicularis* | 130 | 325811 | B80 |
| Infection | Enterovirus | 66 | 325811 | A800, A801, A802, A803, A804, A809, A850, A870, A880, B084, B085, B303, B341, B971, J203, J206, J207 |
| Infection | *Escherichia coli* | 4452 | 325811 | A040, A041, A042, A043, A044, B962, J155 |
| Infection | *Giardia lamblia* | 58 | 325811 | A071 |
| Infection | *Haemophilus influenzae* | 539 | 325811 | A413, A492, B963, J14, J201 |
| Infection | *Helicobacter pylori* | 1888 | 325811 | B980 |
| Infection | Hepatitis A | 92 | 325811 | B150, B159 |
| Infection | Hepatitis C | 453 | 325811 | B171, B182 |
| Infection | HSV | 443 | 325811 | A600, A601, A609, B000, B001, B002, B003, B004, B005, B007, B008, B009 |
| Infection | Influenza | 630 | 325811 | J09, J100, J101, J108, J110, J111, J118 |
| Infection | *Klebsiella pneumoniae* | 698 | 325811 | B961, J150 |
| Infection | *Legionella spp.* | 74 | 325811 | A481, A482 |

| | | | | |
|---|---|---|---|---|
| Infection | Measles | 2 | 325811 | B050, B051, B052, B053, B054, B058, B059 |
| Infection | Molluscum contagiosum virus (MOCV) | 85 | 325811 | B081 |
| Infection | Mumps | 9 | 325811 | B260, B261, B262, B263, B268, B269 |
| Infection | *Neisseria meningitidis* | 76 | 325811 | A390, A391, A392, A393, A394, A395, A398, A399 |
| Infection | Norovirus | 159 | 325811 | A081 |
| Infection | Papillomavirus | 1529 | 325811 | A630, B07, B977 |
| Infection | *Pneumocystis jirovecii* | 80 | 325811 | B59 |
| Infection | Polio | 21 | 325811 | A800, A801, A802, A803, A804, A809 |
| Infection | *Proteus spp.* | 422 | 325811 | B964 |
| Infection | Rubella | 1 | 325811 | B060, B068, B069 |
| Infection | *Salmonella typhi* | 11 | 325811 | A010 |
| Infection | *Staphylococcus aureus* | 3429 | 325811 | A410, B956 |
| Infection | Streptococcus A | 277 | 325811 | A400, B950 |
| Infection | Streptococcus B | 409 | 325811 | A401, B951, J153 |
| Infection | Streptococcus D | 203 | 325811 | A402, B952 |
| Infection | *Streptococcus pneumoniae* | 644 | 325811 | A403, B953, J13, M001 |
| Infection | VZV (Chickenpox) | 96 | 325811 | B010, B011, B012, B018, B019 |
| Infection | VZV (Combined) | 834 | 325811 | B010, B011, B012, B018, B019, B020, B021, B022, B023, B027, B028, B029 |
| Infection | VZV (Zoster) | 741 | 325811 | B020, B021, B022, B023, B027, B028, B029 |
| Inflammation (Other) | Coronary artery disease | 41906 | 355754 | I20, I21, I22, I23, I24, I25 |
| Inflammation (Other) | COPD | 12805 | 384873 | J440, J441, J448, J449 |
| Inflammation (Other) | Diabetes mellitus | 3644 | 394034 | E106, E107, E108, E109 |
| Inflammation (Other) | Encephalomyelitis | 239 | 397439 | G040, G041, G048, G049 |
| Inflammation (Other) | Inflammatory polyarthropathy | 151 | 397527 | M0640, M0641, M0642, M0643, M0644, M0645, M0646, M0647, M0648, M0649 |
| Inflammation (Other) | Motor neuron disease | 347 | 397331 | G122 |
| Inflammation (Chronic) | Multiple sclerosis | 1686 | 395992 | G35 |
| Inflammation (Other) | Myocardial infarction | 16643 | 381017 | I21, I22, I252 |
| Inflammation (Other) | Parkinson's | 1849 | 395829 | G20 |

| | | | | |
|---|---|---|---|---|
| Inflammation (Chronic) | Psoriasis | 3262 | 394416 | L400, L401, L402, L403, L404, L405, L408, L409 |
| Inflammation (Chronic) | Rheumatoid arthritis | 4539 | 393139 | M0500, M0501, M0502, M0503, M0504, M0505, M0506, M0507, M0508, M0509, M0510, M0511, M0512, M0513, M0514, M0515, M0516, M0517, M0518, M0519, M0520, M0521, M0522, M0523, M0524, M0525, M0526, M0527, M0528, M0529, M0530, M0531, M0532, M0533, M0534, M0535, M0536, M0537, M0538, M0539, M0580, M0581, M0582, M0583, M0584, M0585, M0586, M0587, M0589, M0590, M0591, M0592, M0593, M0594, M0595, M0596, M0597, M0598, M0599, M0600, M0601, M0602, M0603, M0604, M0605, M0606, M0607, M0608, M0609, M0610, M0611, M0612, M0613, M0614, M0615, M0616, M0617, M0618, M0619, M0620, M0621, M0622, M0623, M0624, M0625, M0626, M0627, M0628, M0629, M0630, M0631, M0632, M0633, M0634, M0635, M0636, M0637, M0638, M0639, M0640, M0641, M0642, M0643, M0644, M0645, M0646, M0647, M0648, M0649, M0680, M0681, M0682, M0683, M0684, M0685, M0686, M0687, M0688, M0689, M0690, M0691, M0692, M0693, M0694, M0695, M0696, M0697, M0698, M0699 |
| Inflammation (Other) | Rosacea | 439 | 397239 | L710, L711, L718, L719 |
| Inflammation (Chronic) | Sarcoidosis | 826 | 396851 | D860, D861, D862, D863, D868, D869 |
| Inflammation (Chronic) | Systemic lupus erythematosus | 542 | 397136 | M320, M321, M328, M329, M3290 |
| Inflammation (Other) | Ulcerative colitis | 4217 | 393443 | K51 |
| Other | Alzheimer's | 880 | 396798 | G300, G301, G308, G309 |
| Infection | Additional codes included in super-phenotype | NA | NA | A000, A001, A009, A011, A012, A013, A014, A020, A021, A022, A028, A029, A030, A031, A032, A033, A038, A039, A051, A052, A053, A054, A060, A061, A062, A063, A064, A065, A066, A067, A068, A069, A072, A080, A082, A150, A151, A152, A153, A154, A155, A156, A157, A158, A159, A160, A161, A162, A163, A164, A165, A167, A168, A169, A170, A171, A178, A179, A180, A181, A182, A183, A184, A185, A186, A187, A188, A190, A191, A192, A198, A199, A200, A201, A202, A203, A207, A208, A209, A220, A221, A222, A227, A228, A229, A230, A231, A232, A233, A238, A239, A240, A241, A242, A243, A244, A260, A267, A268, A269, A270, A278, A279, A280, A300, A301, A302, A303, A304, A305, A308, A309, A310, A311, A318, A319, A320, A321, A327, A328, A329, |

A33, A34, A35, A371, A378, A379, A38, A408, A409, A420, A421, A422, A427, A428, A429, A440, A441, A448, A449, A46, A480, A484, A490, A491, A493, A510, A511, A512, A513, A514, A515, A519, A520, A521, A522, A523, A527, A528, A529, A530, A539, A55, A560, A561, A562, A563, A564, A568, A57, A58, A590, A598, A599, A65, A660, A661, A662, A663, A664, A665, A666, A667, A668, A669, A670, A671, A672, A673, A679, A680, A681, A689, A692, A70, A710, A711, A719, A740, A748, A749, A750, A751, A752, A753, A759, A770, A771, A772, A773, A778, A779, A78, A790, A791, A820, A821, A829, A830, A831, A832, A833, A834, A836, A840, A841, A848, A849, A851, A871, A872, A90, A91, A920, A921, A922, A923, A932, A950, A951, A959, A970, A971, A972, A979, A981, A982, B03, B04, B080, B083, B160, B161, B162, B169, B170, B172, B178, B179, B180, B181, B188, B189, B190, B199, B200, B201, B202, B203, B204, B205, B206, B207, B208, B209, B210, B211, B212, B213, B217, B218, B219, B220, B221, B222, B227, B230, B231, B232, B238, B24, B300, B301, B331, B333, B340, B342, B343, B350, B351, B352, B353, B354, B355, B356, B358, B359, B360, B361, B362, B363, B368, B369, B380, B381, B382, B383, B384, B387, B388, B389, B390, B391, B392, B393, B394, B395, B399, B400, B401, B402, B403, B407, B408, B409, B410, B417, B418, B419, B420, B421, B427, B428, B429, B430, B431, B432, B438, B439, B450, B451, B452, B453, B457, B458, B459, B460, B461, B462, B463, B464, B465, B468, B469, B470, B471, B479, B480, B481, B482, B483, B484, B487, B488, B49, B510, B518, B519, B520, B528, B529, B530, B531, B538, B550, B551, B552, B559, B560, B561, B569, B570, B571, B572, B573, B574, B575, B580, B581, B582, B583, B588, B589, B601, B650, B651, B652, B653, B658, B659, B663, B670, B671, B672, B673, B674, B675, B676, B677, B678, B679, B680, B681, B689, B770, B778, B779, B780, B781, B787, B789, B832, B850, B851, B852, B86, B954, B955, B957, B958, B960, B965, B966, B967, B970, B973, B974, B975, B976, B981, G000, G002, G003, J020, J030, J122, J123, J151, J152, J154, J200, J202, J204, J205, J210, J211, L00, M000, M002

**Supplementary Table 4:** Phenotype definitions of self-reported phenotypes

| Phenotype group | Phenotype | Cases | Controls | UK Biobank code(s) |
|---|---|---|---|---|
| Allergy | Allergy (unspecified) | 3584 | 369427 | 1374 |
| Allergy | Asthma | 57509 | 315502 | 1111 |
| Allergy | Eczema | 14021 | 358990 | 1452 |
| Allergy | Hayfever | 30537 | 342474 | 1387 |
| Infection | Diphtheria | 81 | 298210 | 1574 |
| Infection | EBV | 1035 | 298210 | 1567 |
| Infection | *Helicobacter pylori* | 1532 | 298210 | 1442 |
| Infection | Hepatitis A | 252 | 298210 | 1578 |
| Infection | Hepatitis C | 135 | 298210 | 1580 |
| Infection | HSV | 260 | 298210 | 1575 |
| Infection | Measles | 5560 | 298210 | 1568 |
| Infection | Mumps | 3384 | 298210 | 1569 |
| Infection | Pertussis | 1282 | 298210 | 1572 |
| Infection | Polio | 402 | 298210 | 1526 |
| Infection | Rubella | 1996 | 298210 | 1570 |
| Infection | Scarlet fever | 1012 | 298210 | 1677 |
| Infection | *Staphylococcus aureus* (MRSA) | 72 | 298210 | 1566 |
| Infection | Tonsillitis | 4893 | 298210 | 1598 |
| Infection | Tuberculosis | 2581 | 298210 | 1440 |
| Infection | Typhoid fever | 60 | 298210 | 1577 |
| Infection | VZV (Chickenpox) | 6886 | 298210 | 1571 |
| Infection | VZV (Combined) | 7475 | 298210 | 1571, 1573, 1674 |
| Infection | VZV (Shingles) | 1067 | 298210 | 1573 |
| Inflammation | Coronary artery disease | 2015 | 370977 | 1082 |
| Inflammation | COPD | 1895 | 371116 | 1112 |
| Inflammation | Diabetes (type 1) | 478 | 372533 | 1222 |
| Inflammation | Multiple sclerosis | 1738 | 371273 | 1261 |
| Inflammation | Myocardial infarction | 11592 | 361400 | 1075 |
| Inflammation | Parkinson's | 901 | 372110 | 1262 |
| Inflammation | Psoriasis | 5861 | 367150 | 1453 |
| Inflammation | Rheumatoid arthritis | 5713 | 367298 | 1464 |
| Inflammation | Rosacea | 1010 | 372001 | 1660 |
| Inflammation | Sarcoidosis | 1030 | 371981 | 1371 |
| Inflammation | Systemic lupus erythematosus | 636 | 372375 | 1381 |
| Inflammation | Ulcerative colitis | 2635 | 370357 | 1463 |
| Other | Abnormal cervical smear | 804 | 201181 | 1663 |
| Other | Alzheimer's | 137 | 372874 | 1263 |
| Other | Cervical abnormality | 1225 | 200760 | 1554, 1663 |
| Other | Cervical intraepithelial neoplasia | 433 | 201552 | 1554 |
| Infection | Additional codes included in super-phenotype | NA | NA | 1156, 1196, 1244, 1246, 1247, 1274, 1412, 1416, 1418, 1439, 1514, 1515, |

| | |
|---|---|
| | 1576, 1579, 1581, 1582, 1594, 1657, 1676, 1678 |

**Supplementary Table 5:** Association statistics for rs28362491 and self-reported and ICD-10 derived phenotypes after exclusion of phenotypes where the regression model was poorly fitted

| Phenotype group | Phenotype | OR | 95% CI | P |
|---|---|---|---|---|
| Allergy | Allergic dermatitis (ICD) | 0.957 | 0.747-1.227 | 0.729 |
| Allergy | Allergy (unspecified) (SR) | 0.986 | 0.940-1.033 | 0.547 |
| Allergy | Asthma | 0.993 | 0.982-1.005 | 0.280 |
| Allergy | Atopic dermatitis (ICD) | 0.850 | 0.637-1.133 | 0.267 |
| Allergy | Eczema (SR) | 0.981 | 0.957-1.005 | 0.123 |
| Allergy | Hayfever | 0.965 | 0.949-0.981 | $3.57\times10^{-5}$ |
| Allergy | Seborrhoeic dermatitis (ICD) | 0.817 | 0.601-1.111 | 0.197 |
| Infection | *Aspergillus spp.* (ICD) | 0.946 | 0.786-1.140 | 0.560 |
| Infection | *Campylobacter spp.* (ICD) | 1.001 | 0.885-1.132 | 0.986 |
| Infection | *Candida spp.* (ICD) | 0.993 | 0.947-1.042 | 0.778 |
| Infection | *Clostridium difficile* (ICD) | 1.018 | 0.930-1.115 | 0.697 |
| Infection | CMV (ICD) | 1.094 | 0.890-1.345 | 0.395 |
| Infection | *Corynebacterium diphtheriae* | 0.912 | 0.667-1.248 | 0.566 |
| Infection | EBV | 1.067 | 0.979-1.162 | 0.143 |
| Infection | *Enterobius vermicularis* (ICD) | 1.110 | 0.868-1.420 | 0.406 |
| Infection | Enterovirus (ICD) | 0.989 | 0.698-1.399 | 0.948 |
| Infection | *Escherichia coli* (ICD) | 1.026 | 0.984-1.071 | 0.230 |
| Infection | *Giardia lamblia* (ICD) | 1.026 | 0.708-1.485 | 0.894 |
| Infection | *Haemophilus influenzae* (ICD) | 1.012 | 0.896-1.143 | 0.850 |
| Infection | *Helicobacter pylori* | 1.022 | 0.974-1.073 | 0.371 |
| Infection | Hepatitis A | 1.027 | 0.882-1.195 | 0.734 |
| Infection | Hepatitis C | 1.122 | 0.983-1.281 | 0.087 |
| Infection | HSV | 1.032 | 0.926-1.149 | 0.570 |
| Infection | Influenza (ICD) | 1.120 | 1.000-1.253 | $4.90\times10^{-2}$ |
| Infection | *Klebsiella pneumoniae* (ICD) | 1.053 | 0.947-1.172 | 0.339 |
| Infection | *Legionella spp*. (ICD) | 1.524 | 1.097-2.118 | $1.21\times10^{-2}$ |
| Infection | Measles | 0.978 | 0.941-1.016 | 0.247 |
| Infection | MOCV (ICD) | 1.056 | 0.778-1.434 | 0.726 |
| Infection | Mumps | 0.996 | 0.948-1.045 | 0.862 |
| Infection | *Neisseria meningitidis* (ICD) | 0.704 | 0.509-0.974 | $3.43\times10^{-2}$ |
| Infection | Norovirus (ICD) | 1.208 | 0.966-1.511 | 0.098 |
| Infection | Papillomavirus (ICD) | 1.095 | 1.019-1.177 | $1.38\times10^{-2}$ |
| Infection | Pertussis | 0.984 | 0.909-1.065 | 0.693 |
| Infection | *Pneumocystis jirovecii* (ICD) | 1.007 | 0.735-1.381 | 0.963 |
| Infection | Polio | 0.958 | 0.835-1.100 | 0.544 |
| Infection | *Proteus spp*. (ICD) | 1.143 | 0.996-1.311 | 0.057 |
| Infection | *Staphylococcus aureus* | 1.012 | 0.964-1.062 | 0.635 |
| Infection | Streptococcus A | 1.072 | 0.991-1.160 | 0.082 |
| Infection | Streptococcus B (ICD) | 1.063 | 0.924-1.222 | 0.391 |
| Infection | Streptococcus D (ICD) | 0.966 | 0.792-1.177 | 0.729 |
| Infection | *Streptococcus pneumoniae* (ICD) | 1.068 | 0.956-1.193 | 0.246 |
| Infection | Tonsillitis (SR) | 0.976 | 0.937-1.016 | 0.235 |

| | | | | |
|---|---|---|---|---|
| Infection | Tuberculosis (SR) | 0.984 | 0.931-1.041 | 0.576 |
| Infection | VZV (Chicken pox) | 0.983 | 0.950-1.017 | 0.313 |
| Infection | VZV (Combined) | 0.994 | 0.963-1.026 | 0.704 |
| Infection | VZV (Shingles) | 1.028 | 0.961-1.099 | 0.421 |
| Inflammation (Other) | Coronary artery disease | 1.004 | 0.989-1.019 | 0.595 |
| Inflammation (Other) | COPD | 1.016 | 0.991-1.042 | 0.206 |
| Inflammation (Other) | Encephalomyelitis (ICD) | 1.048 | 0.874-1.258 | 0.611 |
| Inflammation (Other) | Inflammatory polyarthropathy (ICD) | 0.864 | 0.687-1.087 | 0.213 |
| Inflammation (Other) | Motor neuron disease (ICD) | 1.062 | 0.913-1.236 | 0.435 |
| Inflammation (Chronic) | Multiple Sclerosis | 1.047 | 0.985-1.113 | 0.144 |
| Inflammation (Other) | Myocardial infarction | 1.006 | 0.985-1.028 | 0.550 |
| Inflammation (Other) | Parkinson's | 1.044 | 0.981-1.112 | 0.177 |
| Inflammation (Chronic) | Psoriasis | 1.044 | 1.005-1.083 | $2.55\times10^{-2}$ |
| Inflammation (Chronic) | Rheumatoid Arthritis | 0.986 | 0.955-1.018 | 0.389 |
| Inflammation (Other) | Rosacea | 0.979 | 0.908-1.055 | 0.577 |
| Inflammation (Other) | Ulcerative colitis | 1.079 | 1.037-1.124 | $2.15\times10^{-4}$ |
| Other | Abnormal cervical smear (SR) | 0.990 | 0.896-1.093 | 0.836 |
| Other | Alzheimer's | 1.109 | 1.012-1.215 | $2.65\times10^{-2}$ |
| Other | Cervical abnormality (SR) | 1.055 | 0.973-1.144 | 0.196 |
| Other | Cervical intraepithelial neoplasia (SR) | 1.166 | 1.020-1.334 | $2.48\times10^{-2}$ |
| | Allergy | 0.988 | 0.978-0.998 | $1.84\times10^{-2}$ |
| | Chronic inflammation | 1.020 | 0.998-1.042 | 0.075 |
| | Infection | 1.014 | 1.001-1.027 | $3.31\times10^{-2}$ |

**Supplementary Table 6:** Association statistics for rs28362491 and mortality associated with allergy, infection, or inflammation

| Phenotype | Cases | Controls | OR | 95% CI | P |
|---|---|---|---|---|---|
| Allergy | 131 | 25205 | 0.846 | 0.658-1.088 | 0.192 |
| Infection | 1244 | 24092 | 1.065 | 0.982-1.155 | 0.129 |
| Inflammation | 4421 | 20915 | 1.038 | 0.991-1.087 | 0.117 |

**Supplementary Table 7:** Association statistics for rs28362491 and blood cell counts

| Phenotype | Beta | SE | P |
|---|---|---|---|
| Leukocyte count | $-9.50\times10^{-5}$ | $1.97\times10^{-3}$ | 0.960 |
| Lymphocyte count | $1.98\times10^{-2}$ | $1.95\times10^{-3}$ | $3.30\times10^{-24}$ |
| Basophil count | $-1.25\times10^{-2}$ | $2.08\times10^{-3}$ | $2.00\times10^{-9}$ |
| Eosinophil count | $6.49\times10^{-3}$ | $1.96\times10^{-3}$ | $9.30\times10^{-4}$ |
| Monocyte count | $-1.50\times10^{-2}$ | $1.89\times10^{-3}$ | $2.70\times10^{-15}$ |
| Neutrophil count | $-7.27\times10^{-3}$ | $1.99\times10^{-3}$ | $2.50\times10^{-4}$ |
| RBC count | $-1.04\times10^{-2}$ | $1.67\times10^{-3}$ | $4.80\times10^{-10}$ |
| Platelet count | $-5.79\times10^{-3}$ | $1.79\times10^{-3}$ | $1.30\times10^{-3}$ |

**Supplementary Table 8:** Colocalisation results for GWAS and eQTL phenotype pairs

| Phenotype 1 | Phenotype 2 | Lead SNP 1 | Lead SNP 2 | PP H0 | PP H1 | PP H2 | PP H3 | PP H4 |
|---|---|---|---|---|---|---|---|---|
| EBV (EA-D) | EBV (ZEBRA) | rs230493 | rs4648058 | $2.02\times10^{-4}$ | $3.31\times10^{-3}$ | $1.01\times10^{-2}$ | $1.64\times10^{-1}$ | $8.22\times10^{-1}$ |
| EBV (ZEBRA) | HHV-6 (IE1A) | rs4648058 | rs1598859 | $1.28\times10^{-8}$ | $6.42\times10^{-7}$ | $2.88\times10^{-3}$ | $1.43\times10^{-1}$ | $8.54\times10^{-1}$ |
| HHV-6 (IE1A) | HHV-7 (U14) | rs1598859 | rs4648052 | $9.53\times10^{-8}$ | $2.15\times10^{-2}$ | $1.82\times10^{-6}$ | $4.09\times10^{-1}$ | $5.69\times10^{-1}$ |
| HHV-7 (U14) | HIV (Env) | rs4648052 | rs10013613 | $4.71\times10^{-4}$ | $9.01\times10^{-3}$ | $6.80\times10^{-3}$ | $1.28\times10^{-1}$ | $8.55\times10^{-1}$ |
| HIV (Env) | HPV18 (L1) | rs10013613 | rs4648052 | $9.18\times10^{-4}$ | $1.33\times10^{-2}$ | $9.37\times10^{-3}$ | $1.34\times10^{-1}$ | $8.43\times10^{-1}$ |
| HPV18 (L1) | HTLV-1 (Env) | rs4648052 | rs4648058 | $5.38\times10^{-4}$ | $5.49\times10^{-3}$ | $1.31\times10^{-2}$ | $1.32\times10^{-1}$ | $8.49\times10^{-1}$ |
| HTLV-1 (Env) | KSHV (K8.1) | rs4648058 | rs10013613 | $7.84\times10^{-5}$ | $1.91\times10^{-3}$ | $9.39\times10^{-3}$ | $2.27\times10^{-1}$ | $7.61\times10^{-1}$ |
| KSHV (K8.1) | *T. gondii* (sag1) | rs10013613 | rs10013613 | $7.81\times10^{-5}$ | $9.36\times10^{-3}$ | $8.16\times10^{-4}$ | $9.59\times10^{-2}$ | $8.94\times10^{-1}$ |
| HHV-6 (IE1A) | Basophil count | rs1598859 | rs2168805 | $1.33\times10^{-10}$ | $2.99\times10^{-5}$ | $1.70\times10^{-6}$ | $3.81\times10^{-1}$ | $6.19\times10^{-1}$ |
| HHV-6 (IE1A) | Eosinophil count | rs1598859 | rs230507 | $3.71\times10^{-21}$ | $8.35\times10^{-16}$ | $6.10\times10^{-7}$ | $1.36\times10^{-1}$ | $8.64\times10^{-1}$ |
| HHV-6 (IE1A) | Lymphocyte count | rs1598859 | rs35680095 | $7.00\times10^{-18}$ | $1.58\times10^{-12}$ | $4.44\times10^{-6}$ | $9.99\times10^{-1}$ | $1.45\times10^{-7}$ |
| HHV-6 (IE1A) | Monocyte count | rs1598859 | rs980455 | $4.04\times10^{-17}$ | $9.09\times10^{-12}$ | $1.33\times10^{-6}$ | $2.97\times10^{-1}$ | $7.03\times10^{-1}$ |
| HHV-6 (IE1A) | Red blood cell count | rs1598859 | rs230539 | $3.15\times10^{-12}$ | $7.08\times10^{-7}$ | $9.36\times10^{-7}$ | $2.09\times10^{-1}$ | $7.91\times10^{-1}$ |
| HHV-6 (IE1A) | Platelet count | rs1598859 | rs62328536 | $2.49\times10^{-6}$ | $5.60\times10^{-1}$ | $4.75\times10^{-7}$ | $1.06\times10^{-1}$ | $3.33\times10^{-1}$ |
| HHV-6 (IE1A) | eQTL: Monocytes (unstim) | rs1598859 | rs72696119 | $5.34\times10^{-7}$ | $1.20\times10^{-1}$ | $1.26\times10^{-6}$ | $2.82\times10^{-1}$ | $5.98\times10^{-1}$ |
| HHV-6 (IE1A) | eQTL: Monocytes (LPS2) | rs1598859 | rs4698856 | $1.39\times10^{-8}$ | $3.14\times10^{-3}$ | $1.53\times10^{-6}$ | $3.43\times10^{-1}$ | $6.53\times10^{-1}$ |
| HHV-6 (IE1A) | eQTL: Monocytes (LPS24) | rs1598859 | rs28882677 | $1.60\times10^{-8}$ | $3.61\times10^{-3}$ | $2.10\times10^{-6}$ | $4.72\times10^{-1}$ | $5.25\times10^{-1}$ |
| HHV-6 (IE1A) | eQTL: Monocytes (IFN) | rs1598859 | rs4648055 | $9.37\times10^{-12}$ | $2.11\times10^{-6}$ | $6.76\times10^{-7}$ | $1.50\times10^{-1}$ | $8.50\times10^{-1}$ |
| HHV-6 (IE1A) | eQTL: Neutrophils | rs1598859 | rs4698857 | $6.31\times10^{-8}$ | $1.42\times10^{-2}$ | $7.72\times10^{-7}$ | $1.72\times10^{-1}$ | $8.14\times10^{-1}$ |
| HHV-6 (IE1A) | eQTL: Natural killer cells | rs1598859 | rs72696119 | $3.90\times10^{-8}$ | $8.78\times10^{-3}$ | $1.18\times10^{-6}$ | $2.63\times10^{-1}$ | $7.28\times10^{-1}$ |
| Monocyte count | eQTL: Monocytes (unstim) | rs980455 | rs72696119 | $1.44\times10^{-12}$ | $4.73\times10^{-2}$ | $3.40\times10^{-12}$ | $1.10\times10^{-1}$ | $8.43\times10^{-1}$ |
| Monocyte count | eQTL: Monocytes (LPS2) | rs980455 | rs4698856 | $2.75\times10^{-14}$ | $9.03\times10^{-4}$ | $3.02\times10^{-12}$ | $9.74\times10^{-2}$ | $9.02\times10^{-1}$ |
| Monocyte count | eQTL: Monocytes (LPS24) | rs980455 | rs28882677 | $2.15\times10^{-14}$ | $7.05\times10^{-4}$ | $2.81\times10^{-12}$ | $9.05\times10^{-2}$ | $9.09\times10^{-1}$ |
| Monocyte count | eQTL: Monocytes (IFN) | rs980455 | rs4648055 | $1.44\times10^{-16}$ | $4.73\times10^{-6}$ | $1.04\times10^{-11}$ | $3.40\times10^{-1}$ | $6.60\times10^{-1}$ |
| Neutrophil count | eQTL: Neutrophils | rs2903281 | rs4698857 | $2.33\times10^{-3}$ | $3.22\times10^{-2}$ | $2.84\times10^{-2}$ | $3.93\times10^{-1}$ | $5.44\times10^{-1}$ |
| Lymphocyte count | eQTL: Natural killer cells | rs35680095 | rs72696119 | $5.05\times10^{-14}$ | $3.20\times10^{-2}$ | $1.52\times10^{-12}$ | $9.65\times10^{-1}$ | $2.78\times10^{-3}$ |

*PP: Posterior probability

**References**


1. Brenner, N. *et al.* Validation of Multiplex Serology detecting human herpesviruses 1-5. *PLoS One* **13**, e0209379 (2018).
2. Brenner, N. *et al.* Validation of Multiplex Serology for human hepatitis viruses B and C, human T-lymphotropic virus 1 and Toxoplasma gondii. *PLoS One* **14**, e0210407 (2019).
3. Dondog, B. *et al.* Hepatitis C Virus Seroprevalence in Mongolian Women Assessed by a Novel Multiplex Antibody Detection Assay. *Cancer Epidemiology Biomarkers & Prevention* **24**, 1360–1365 (2015).
4. Kranz, L. M. *et al.* Development and validation of HIV-1 Multiplex Serology. *Journal of Immunological Methods* **466**, 47–51 (2019).
5. Kjærheim, K. *et al.* Absence of SV40 antibodies or DNA fragments in prediagnostic mesothelioma serum samples. *International Journal of Cancer* **120**, 2459–2465 (2007).
6. Gossai, A. *et al.* Prospective Study of Human Polyomaviruses and Risk of Cutaneous Squamous Cell Carcinoma in the United States. *Cancer Epidemiology Biomarkers & Prevention* **25**, 736–744 (2016).
7. Robles, C. *et al.* Seroreactivity against Merkel cell polyomavirus and other polyomaviruses in chronic lymphocytic leukaemia, the MCC-Spain study. *Journal of General Virology* **96**, 2286–2292 (2015).
8. Sehr, P., Zumbach, K. & Pawlita, M. A generic capture ELISA for recombinant proteins fused to glutathione S-transferase: validation for HPV serology. *Journal of Immunological Methods* **253**, 153–162 (2001).
9. Sehr, P., Müller, M., Höpfl, R., Widschwendter, A. & Pawlita, M. HPV antibody detection by ELISA with capsid protein L1 fused to glutathione S-transferase. *Journal of Virological Methods* **106**, 61–70 (2002).
10. Hulstein, S. H. *et al.* Differences in Chlamydia trachomatis seroprevalence between ethnic groups cannot be fully explained by socioeconomic status, sexual healthcare seeking behavior or sexual risk behavior: a cross-sectional analysis in the HEalthy LIfe in an Urban Setting (HELIUS) study. *BMC Infectious Diseases* **18**, 612 (2018).
11. Trabert, B. *et al.* Antibodies Against Chlamydia trachomatis and Ovarian Cancer Risk in Two Independent Populations. *JNCI: Journal of the National Cancer Institute* **111**, 129–136 (2018).
12. Michel, A., Waterboer, T., Kist, M. & Pawlita, M. Helicobacter pylori Multiplex Serology. *Helicobacter* **14**, 525–535 (2009).
