## Extended data for "A frequent ancestral NFKB1 variant predicts risk of infection or allergy"

**Extended Data Table 1:** Top variants for genomic regions excluding the extended MHC region associated with antibody responses to 45 pathogen-derived antigens. Betas are reported relative to the minor allele.

| Chr | Region | Antigen | SNP | MAF | Beta | SE | P |
|---|---|---|---|---|---|---|---|
| 3 | 68638922-69293121 | *C. trachomatis* (mompA) | 3:68985092_TAGTGACA_T | 0.030 | 0.279 | 0.045 | $6.50\times10^{-10}$ |
| | | *C. trachomatis* (mompD) | 3:68985092_TAGTGACA_T | 0.030 | 0.262 | 0.045 | $7.10\times10^{-9}$ |
| 4 | 103390496-104199922 | EBV (EA-D) | rs230493 | 0.355 | -0.068 | 0.015 | $3.60\times10^{-6}$ |
| | | EBV (VCAp18) | rs1289231 | 0.077 | 0.126 | 0.028 | $7.00\times10^{-6}$ |
| | | EBV (ZEBRA) | rs3755867 | 0.332 | -0.076 | 0.015 | $3.60\times10^{-7}$ |
| | | EBV (ZEBRA) | rs4699031 | 0.332 | -0.076 | 0.015 | $3.60\times10^{-7}$ |
| | | HHV-6 (IE1A) | 4:103406914_TA_T | 0.366 | -0.096 | 0.015 | $1.30\times10^{-10}$ |
| | | HHV-7 (U14) | rs4648052 | 0.375 | -0.072 | 0.014 | $5.70\times10^{-7}$ |
| | | HIV (env) | rs567727578 | 0.468 | -0.068 | 0.015 | $4.00\times10^{-6}$ |
| | | HPV18 (L1) | rs4648052 | 0.375 | -0.068 | 0.015 | $3.90\times10^{-6}$ |
| | | HTLV-1 (env) | rs4648068 | 0.330 | -0.075 | 0.015 | $1.00\times10^{-6}$ |
| | | HTLV-1 (env) | rs4648058 | 0.330 | -0.075 | 0.015 | $1.00\times10^{-6}$ |
| | | KSHV (K8.1) | rs10013613 | 0.403 | -0.074 | 0.014 | $2.50\times10^{-7}$ |
| | | KSHV (K8.1) | rs74462352 | 0.397 | -0.075 | 0.014 | $2.50\times10^{-7}$ |
| | | *T. gondii* (sag1) | rs10013613 | 0.403 | -0.067 | 0.014 | $3.20\times10^{-6}$ |
| 5 | 138114233-139053851 | MCV (VP1) | rs13181561 | 0.265 | 0.155 | 0.016 | $4.90\times10^{-22}$ |
| 7 | 150011299-150484264 | HTLV-1 (gag) | rs12534190 | 0.230 | 0.166 | 0.017 | $9.90\times10^{-23}$ |
| 11 | 118762073-118811315 | HHV-7 (U14) | rs75438046 | 0.028 | -0.278 | 0.043 | $9.80\times10^{-11}$ |
| 14 | 106624768-107228049 | HCV (NS3) | rs75801933 | 0.066 | 0.145 | 0.029 | $6.80\times10^{-7}$ |
| | | HIV (env) | rs4977158 | 0.084 | 0.163 | 0.036 | $4.80\times10^{-6}$ |
| | | HIV (gag) | rs10150853 | 0.099 | 0.140 | 0.030 | $2.30\times10^{-6}$ |
| | | HIV (gag) | rs10138691 | 0.099 | 0.140 | 0.030 | $2.30\times10^{-6}$ |
| | | HPV16 (E6) | rs2337939 | 0.067 | 0.191 | 0.030 | $1.60\times10^{-10}$ |
| | | HPV16 (E7) | rs199944736 | 0.150 | -0.113 | 0.021 | $5.90\times10^{-8}$ |
| | | HPV16 (E7) | rs201829121 | 0.150 | -0.113 | 0.021 | $5.90\times10^{-8}$ |
| | | HPV16 (L1) | rs4774155 | 0.070 | 0.136 | 0.028 | $1.60\times10^{-6}$ |
| | | HPV18 (L1) | rs75801933 | 0.066 | 0.134 | 0.029 | $4.10\times10^{-6}$ |
| | | HTLV-1 (env) | rs75801933 | 0.066 | 0.154 | 0.029 | $1.40\times10^{-7}$ |
| | | *T. gondii* (p22) | rs4977158 | 0.084 | 0.176 | 0.036 | $7.70\times10^{-7}$ |
| | | *T. gondii* (sag1) | rs4977158 | 0.084 | 0.213 | 0.035 | $1.70\times10^{-9}$ |
| 19 | 49097126-49282803 | BKV (VP1) | rs681343 | 0.489 | -0.123 | 0.014 | $3.80\times10^{-18}$ |
| | | JCV (VP1) | rs681343 | 0.489 | -0.134 | 0.014 | $1.70\times10^{-21}$ |
| 22 | 23154058-23220565 | *H. pylori* (UreA) | 22:23169419_GT_G | 0.173 | -0.119 | 0.019 | $9.00\times10^{-10}$ |
| 22 | 41424903-42698602 | HHV-7 (U14) | rs35211694 | 0.245 | 0.117 | 0.019 | $4.40\times10^{-10}$ |

**Extended Data Table 2:** Top variants within the extended MHC region (chr6:25,384,361-34,366,455) associated with antibody responses to 45 pathogen-derived antigens. Betas are reported relative to the minor allele.

| Antigen | SNP | MAF | Beta | SE | P |
|---|---|---|---|---|---|
| BKV (VP1) | rs4713573 | 0.324 | 0.079 | 0.016 | $4.70\times10^{-7}$ |
| CMV (pp150) | rs9275015 | 0.300 | -0.086 | 0.017 | $2.30\times10^{-7}$ |
| CMV (pp52) | rs9270798 | 0.232 | 0.096 | 0.017 | $1.00\times10^{-8}$ |
| *C. trachomatis* (mompA) | rs71536538 | 0.273 | -0.081 | 0.017 | $3.50\times10^{-6}$ |
| *C. trachomatis* (mompD) | rs4367411 | 0.201 | -0.098 | 0.019 | $1.80\times10^{-7}$ |
| *C. trachomatis* (TarpDF2) | rs112438356 | 0.098 | -0.139 | 0.026 | $5.00\times10^{-8}$ |
| *C. trachomatis* (TarpDF2) | rs112327275 | 0.098 | -0.139 | 0.026 | $5.00\times10^{-8}$ |
| EBV (EA-D) | rs2395192 | 0.460 | -0.163 | 0.014 | $3.40\times10^{-31}$ |
| EBV (EBNA1) | rs530423766 | 0.433 | -0.324 | 0.015 | $6.90\times10^{-97}$ |
| EBV (VCAp18) | rs9264759 | 0.208 | 0.158 | 0.018 | $4.00\times10^{-19}$ |
| EBV (ZEBRA) | rs9274704 | 0.278 | 0.306 | 0.016 | $2.70\times10^{-86}$ |
| HBV (HBc) | rs112275579 | 0.365 | 0.079 | 0.017 | $3.80\times10^{-6}$ |
| HBV (HBe) | rs9266092 | 0.391 | 0.070 | 0.015 | $4.90\times10^{-6}$ |
| HCV (NS3) | rs3997848 | 0.323 | 0.082 | 0.015 | $1.20\times10^{-7}$ |
| HHV-6 (IE1A) | rs9265967 | 0.139 | -0.134 | 0.021 | $2.30\times10^{-10}$ |
| HHV-6 (IE1B) | rs28383304 | 0.145 | 0.146 | 0.021 | $1.30\times10^{-12}$ |
| HHV-6 (p101k) | rs4959072 | 0.219 | 0.120 | 0.017 | $5.40\times10^{-12}$ |
| HHV-7 (U14) | rs9270140 | 0.143 | -0.163 | 0.021 | $6.20\times10^{-15}$ |
| HIV (env) | rs41288885 | 0.059 | 0.153 | 0.031 | $1.20\times10^{-6}$ |
| HIV (env) | 6:32748889_CT_C | 0.046 | 0.185 | 0.038 | $1.20\times10^{-6}$ |
| HIV (gag) | rs3134974 | 0.445 | -0.072 | 0.015 | $2.10\times10^{-6}$ |
| HPV16 (E6) | rs35445446 | 0.168 | 0.087 | 0.019 | $6.60\times10^{-6}$ |
| HPV16 (E7) | rs78274956 | 0.495 | -0.130 | 0.016 | $1.20\times10^{-16}$ |
| HPV16 (L1) | rs548129848 | 0.417 | 0.088 | 0.017 | $1.00\times10^{-7}$ |
| HPV18 (L1) | rs6930081 | 0.468 | -0.066 | 0.015 | $5.60\times10^{-6}$ |
| HPV18 (L1) | rs6929819 | 0.468 | -0.066 | 0.014 | $5.60\times10^{-6}$ |
| HSV-1 (1gG) | rs116135626 | 0.187 | 0.151 | 0.025 | $1.20\times10^{-9}$ |
| HSV-2 (2mgG) | rs36152293 | 0.337 | 0.107 | 0.018 | $4.60\times10^{-9}$ |
| HTLV-1 (env) | rs115675626 | 0.088 | -0.154 | 0.027 | $2.20\times10^{-8}$ |
| HTLV-1 (gag | rs9271727 | 0.252 | 0.164 | 0.017 | $2.20\times10^{-22}$ |
| *H. pylori* (CagA) | rs71534592 | 0.121 | -0.172 | 0.032 | $5.60\times10^{-8}$ |
| *H. pylori* (Catalase) | rs9270641 | 0.199 | 0.104 | 0.020 | $3.90\times10^{-7}$ |
| *H. pylori* (HP1564) | rs9268541 | 0.058 | 0.146 | 0.030 | $1.30\times10^{-6}$ |
| *H. pylori* (VacA) | rs35186928 | 0.370 | -0.075 | 0.015 | $3.30\times10^{-7}$ |
| JCV (VP1) | rs71538510 | 0.173 | -0.323 | 0.019 | $8.50\times10^{-66}$ |
| KSHV (K8.1) | rs2844535 | 0.274 | -0.076 | 0.016 | $1.80\times10^{-6}$ |
| MCV (VP1) | rs9269771 | 0.274 | -0.249 | 0.017 | $2.70\times10^{-48}$ |
| *T. gondii* (sag1) | rs144947706 | 0.369 | -0.113 | 0.016 | $1.40\times10^{-12}$ |
| VZV (gE/gI) | rs9273325 | 0.174 | 0.221 | 0.019 | $7.10\times10^{-33}$ |

**Extended Data Table 3**: Published associations between *NFKB1* and disease-related traits

| Published trait | Trait type | Ancestry | SNP | Study size (cases) | P | OR (del) | 95% LCI | 95% UCI | Reference |
|---|---|---|---|---|---|---|---|---|---|
| Allergic rhinitis | Allergy | Caucasian | rs12509403 | 891,367 (120,482) | $1.17\times10^{-15}$ | 0.960 | 0.950 | 0.97 | 57 |
| Allergic sensitisation | Allergy | Caucasian | rs4648050 | 24,481 (8,040) | $2.03\times10^{-8}$ | 0.880 | 0.840 | 0.92 | 57 |
| Combined hayfever and eczema | Allergy | UKB | rs230507 | 346,545 (106,772) | $1.04\times10^{-9}$ | 0.963 | 0.951 | 0.974 | 58 |
| Tonsillectomy | Infection | European | rs230523 | 173,421 (60,098) | $4.54\times10^{-14}$ | 1.070 | 1.060 | 1.08 | 11 |
| Acute respiratory distress syndrome (<65yrs) | Inflammation | European | rs28362491 | NR | $3.00\times10^{-2}$ | 0.106 | 0.014 | 0.825 | 59 |
| Ankylosing spondylitis | Inflammation | European | rs3774937 | 86,475 (52,262) | $7.83\times10^{-10}$ | 1.120 | 1.080 | 1.16 | 60 |
| Behçet's disease | Inflammation | Turkish | rs28362491 | 270 (89) | $4.00\times10^{-3}$ | 0.556 | 0.380 | 0.811 | 61 |
| Coronary artery disease | Inflammation | European | rs28362491 | 220 (120) | $1.50\times10^{-2}$ | 2.880 | 1.210 | 6.84 | 62 |
| Coronary artery disease | Inflammation | India | rs28362491 | 830 (600) | $2.90\times10^{-2}$ | 1.260 | 1.030 | 1.55 | 32 |
| Coronary artery disease | Inflammation | Uygur | rs28362491 | 2020 (960) | <0.001 | 1.581 | 1.222 | 2.046 | 63 |
| Left ventricular dysfunction in CAD* | Inflammation | India | rs28362491 | 600 (190) | $7.00\times10^{-3}$ | 2.340 | 1.310 | 4.17 | 32 |
| Lung injury score in ARDS* | Inflammation | European | rs28362491 | 103 (77) | $3.00\times10^{-4}$ | 3.700 | 1.800 | 7.9 | 64 |
| Mouth Ulcers | Inflammation | European | rs4699030 | 816,850 (145,377) | $5.91\times10^{-11}$ | 1.030 | 1.020 | 1.04 | 65 |
| Myocardial infarction | Inflammation | Caucasian | rs28362491 | 253 (86) | <0.001 | 0.304 | 0.177 | 0.522 | 66 |
| Primary biliary cholangitis | Inflammation | Japanese | rs230534 | 2,886 (1,381) | $1.50\times10^{-7}$ | 1.340 | 1.200 | 1.49 | 67 |
| Primary sclerosing cholangitis | Inflammation | European | rs3774937 | 37,621 (3,408) | $6.11\times10^{-9}$ | 1.167 | 1.107 | 1.229 | 60 |
| Systemic sclerosis | Inflammation | Iran/Turkey | rs4648133 | 2107 (764) | $3.11\times10^{-7}$ | 1.470 | 1.270 | 1.7 | 68 |
| TR schizophrenia* | Inflammation | Han Chinese | rs230529 | 1,610 (804) | $1.74\times10^{-7}$ | 1.450 | 1.260 | 1.66 | 69 |
| Ulcerative colitis | Inflammation | European | rs3774959 | 47,560 (17,865) | $3.66\times10^{-12}$ | 1.118 | 1.077 | 1.159 | 70 |
| Ulcerative colitis | Inflammation | European | rs3774937 | 48,626 (14,413) | $1.33\times10^{-11}$ | 1.107 | 1.075 | 1.142 | 60 |
| Ulcerative colitis | Inflammation | European | rs28362491 | 1,152 (350) | $4.30\times10^{-3}$ | 1.570 | 1.140 | 2.16 | 25 |
| Ulcerative colitis | Inflammation | European | rs28362491 | 282 (127) | $1.10\times10^{-2}$ | 2.510 | 1.280 | 4.92 | 71 |
| Cancer risk | Other | Causcasian/Chinese | rs28362491 | 43,000 (18222) | $2.00\times10^{-3}$ | 0.890 | 0.830 | 0.96 | 72 |
| **Published trait** | | **Ancestry** | **SNP** | **Study size (cases)** | **P** | **Beta (del)** | **SE** | | **Reference** |
| Albumin/Globulin Ratio | Other | Japanese | rs60371688 | 98,626 | $1.49\times10^{-12}$ | 0.036 | 0.005 | | 73 |
| Asthma | Allergy | UKB | rs59123962 | 458,699 | $7.50\times10^{-4}$ | -0.002 | 0.001 | | 39 |
| Combined hayfever, allergic rhinitis, and eczema | Allergy | UKB | rs230504 | 458,699 | $6.90\times10^{-13}$ | -0.007 | 0.001 | | 39 |
| Non-albumin protein | Other | Japanese | rs1585213 | 98,538 | $1.47\times10^{-15}$ | -0.037 | 0.005 | | 73 |
| Immunogobulin levels ((A + G)/M) | Other | European | rs4648052 | 19,219 | $3.00\times10^{-7}$ | -0.060 | NR | | 13 |
| Immunogobulin levels (A - M) | Other | European | rs4648052 | 19,219 | $2.30\times10^{-7}$ | -0.070 | NR | | 13 |

*ARDS: Acute respiratory distress syndrome; CAD: Coronary artery disease; TR: Treatment resistant

**Extended Data Table 4:** Top variants associated with antibody response in the UK Biobank and CoLaus datasets following meta-analysis

| Antigen | SNP ID (UKB) | UK Biobank | | | CoLaus | | | Meta-analysis (fixed-effects) | | |
|---|---|---|---|---|---|---|---|---|---|---|
| | | beta | SE | P | beta | SE | P | beta | SE | P |
| EBV (EAD) | rs2395192 | -0.163 | 0.014 | $3.40\times10^{-31}$ | -0.111 | 0.027 | $4.83\times10^{-5}$ | -0.152 | 0.012 | $3.54\times10^{-34}$ |
| EBV (ZEBRA) | rs9274728 | -0.306 | 0.016 | $3.70\times10^{-86}$ | -0.323 | 0.032 | $1.83\times10^{-24}$ | -0.309 | 0.014 | $8.74\times10^{-109}$ |
| HHV6 (IE1A) | rs8192589 | 0.126 | 0.020 | $4.70\times10^{-10}$ | 0.092 | 0.037 | $1.41\times10^{-2}$ | 0.118 | 0.018 | $2.98\times10^{-11}$ |
| HHV7 (U14) | 6:32593225_CAG_C | 0.198 | 0.026 | $7.20\times10^{-14}$ | 0.218 | 0.044 | $5.77\times10^{-7}$ | 0.203 | 0.023 | $2.44\times10^{-19}$ |
| KSHV (K8.1) | rs2372093 | -0.080 | 0.018 | $1.20\times10^{-5}$ | -0.082 | 0.029 | $5.46\times10^{-3}$ | -0.080 | 0.016 | $2.10\times10^{-7}$ |
| *T. gondii* (sag1) | rs7754119 | 0.103 | 0.015 | $1.10\times10^{-11}$ | 0.053 | 0.023 | $2.04\times10^{-2}$ | 0.088 | 0.013 | $3.95\times10^{-12}$ |
